# A Meta-Analytical Review of Executive Function Skills in Adults who Stutter

**DOI:** 10.1101/2025.09.02.25334917

**Authors:** Levi C. Ofoe, Katerina Ntourou, Shari Clifton, Geoffrey A. Coalson

**Author notes:** Correspondence concerning this article should be addressed to Levi C. Ofoe, Department of Counseling, Higher Education, and Speech-Language Pathology, University of West Georgia, Carrollton, GA 30117. Authors Note: Levi C. Ofoe*, Department of Counseling, Higher Education, and Speech-Language Pathology, University of West Georgia; Katerina Ntourou, Department of Psychological Sciences, Case Western Reserve University; Shari Clifton, Reference & Instructional Services, University of Oklahoma Health Sciences Center; Geoffrey A. Coalson, Moody College of Communication, The University of Texas at Austin. **Conflict of Interest:** The authors report no conflicts of interest.

## Abstract

**Purpose:** Executive function has been identified as a potential area of vulnerability in individuals who stutter. The present study identified and analyzed the data across empirical studies of the executive function skills of adults who do (AWS) and do not stutter (AWNS).

**Method:** Electronic databases, literature reviews, and reference sections of articles and dissertations were searched to identify candidate studies that examined behavioral measures of working memory, inhibition, and/or cognitive flexibility. A total of 39 studies met the eligibility criteria for this meta-analysis. A random-effects model was applied to estimate the pooled effect sizes (Hedges’ *g*) and 95% confidence intervals.

**Results:** AWS were significantly less accurate than AWNS on measures of working memory (Hedges’ *g* = - 0.41, *p* < .001), including nonword repetition (Hedges’ *g* = -.57, *p* < .001), forward digit span (Hedges’ *g* = −0.23, *p* = .02), backward digit span (Hedges’ *g* = -.38, *p* = .004) and operation span tasks (Hedges’ *g* = - .37, *p* = .018). AWS performed comparably to AWNS on inhibition measures (Hedges’ *g* = −0.10, *p* = .37). An insufficient number of published studies were available to conduct a meaningful analysis of cognitive flexibility.

**Conclusions:** Present findings suggest that AWS, as a group, exhibit weaknesses in one component of executive function – working memory – compared to AWNS. Additional research is necessary to determine potential differences in inhibition and cognitive flexibility in AWS.

---

Executive function is a comprehensive term used to describe the general-purpose cognitive processes that underlie goal-directed thoughts and actions. Executive function comprises three core components: working memory, inhibition, and cognitive flexibility (Diamond, 2013; Miyake et al., 2000). Executive function skills emerge during infancy and continue to develop into adolescence and early adulthood (Diamond, 1988; Ferguson et al., 2021). Executive function skills are associated with psychosocial development, mental health, and overall quality of life across the lifespan (Brown & Landgraf, 2010; Carlson, 2005; Garon et al., 2008; Gathercole et al., 2004; Miller et al., 2011). Over the past two decades, more than 35 individual studies have examined the executive function skills of children who do and do not stutter, yielding mixed results (Anderson & Ofoe, 2019). Based on a meta-analytic review, which has more power to statistically detect between-group differences and provide a more accurate summary and direction of effects when compared to individual studies, Ofoe et al. (2018) found that executive function skills in young children who stutter, on average, are less robust than those of their typically fluent peers in two main areas: (1) experimental/behavioral measures of phonological and verbal short-term memory and (2) parent-report questionnaires/measures on inhibition. Findings from more recent studies suggest that the observed weaknesses in executive function skills in children who stutter may persist beyond the preschool-age years (e.g., Eichorn & Pirutinsky, 2021; Paphiti & Eggers, 2022).

According to the *Executive Function Model of Developmental Stuttering* (Anderson & Ofoe, 2019), there are multiple connections between executive function and stuttering. One such link is that weaknesses in executive function skills may directly contribute to the development and/or maintenance of stuttering. In a reciprocal way, the frequent breakdown in the fluency of language production over a period of time could also directly weaken executive function skills. An alternative trajectory, in an indirect manner, involves the intertwined relationship between the executive function skills and language processes, such that weaknesses in one may directly affect the other, and subsequently affect the fluency of language production. Along this latter route, the connection between executive function and stuttering is through language processes (that is, deficits in executive function skills may weaken language skills, which in turn may affect fluency skills). However, other researchers have also suggested potential links between more specific aspects of executive function and (dis)fluent speech production (i.e., working memory: Eichorn et al., 2016; inhibition: Neef et al., 2018; cognitive flexibility: Eichorn et al., 2018; Eichorn & Pirutinsky, 2021), albeit inconsistently (for review, see Anderson & Ofoe, 2019).

Theories regarding the role of executive function and the instances of stuttering are varied, including (but are not limited to) difficulty storing and/or identifying inaccurate phonological information in working memory (Bajaj, 2007; Eichorn et al., 2016), difficulty suppressing an incorrect speech plan before it occurs (e.g., Anderson & Wagovich, 2017), and difficulty revising motoric plans after an error is encountered (e.g., Eggers et al., 2013; Eichorn et al., 2018). Beyond speech production, executive function skills in adults have also been hypothesized to underlie potential cognitive and affective consequences of stuttering (e.g., Coalson et al., 2025; Usler, 2022).

To date, no meta-analytic analysis has been conducted on the differences in executive function skills between adults who do (AWS) and do not stutter (AWNS). Similar to children who stutter, AWS performance on behavioral measures of executive function relative to AWNS are mixed. Such cross-study discrepancies in the adult stuttering literature may be due to methodological inconsistencies (e.g., task selection, sample size, outcome measures). In light of this, the goal of the current study is to systematically identify, synthesize, and analyze the quantitative data across individual studies that have examined the executive function skills of AWS to assess potential between-group differences, if present, relative to AWNS. To provide a contextual framework for the study, we will first discuss the three core components of executive function—working memory, inhibition, and cognitive flexibility—and then briefly review the existing literature on executive function skills in AWS.

## Working Memory

The first core executive function skill is working memory. First proposed as a concept by Miller et al. (1960), working memory refers to a multicomponent, capacity-limited system responsible for temporarily storing (i.e., short-term memory) and manipulating information for complex cognitive purposes (i.e., working memory; Baddeley, 2000; Baddeley & Hitch, 1974; Diamond, 2013). As a resource-limited system, its *short-term memory* component refers to the cognitive capacity for holding information in mind for a brief period (Cowan, 2008), whereas the *working memory* component involves manipulating and updating the current information in mind (Baddeley, 2012). While there are at least ten (10) different models of working memory (see Cowan, 2017; Shah & Miyake, 1999), Baddeley and Hitch’s (1974) model remains the most prominent (D’Esposito & Postle, 2015) and consists of four distinct but interrelated components (a) the phonological loop (or a domain-specific verbal short-term memory), (b) visuospatial sketchpad (or domain-specific visual-spatial short-term memory), (c) the episodic buffer (a system that integrates the verbal and visuospatial information), and (d) the central executive (the attentional command center over processing, manipulation, and recall of verbal and visuospatial information; cf. Baddeley & Hitch, 1994). Thus, Baddeley and Hitch’s model of working memory was selected for our meta-analytical review, given that it is a modular framework compared to the other models (Cowan, 2008; Engle, 2002; Oberauer, 2019).

One component of Baddeley and Hitch’s working memory model, the phonological loop, consists of a store, which holds information for a limited amount of time (typically about two seconds), and an embedded rehearsal mechanism, which refreshes the information in the store to prevent decay (Baddeley, 2004; Baddeley et al., 1998). Researchers have examined the phonological store of short-term memory using a variety of *simple* span tasks. One such task used to measure short-term memory is the *nonword repetition* task (Estes et al., 2007; Gupta, 2003), which involves participants hearing and accurately repeating a string of non-lexical ‘words’ of increasing length and complexity, typically from one syllable (e.g., *jup*) to seven syllables or more (e.g., *dookershatupietazawm*; Wagner et al., 2013).

Another type of span task is the *Forward digit span* task, which is similar to nonword repetition but requires participants to repeat progressively longer series of digits in the correct order immediately after presentation (e.g., Dempster, 1981; Engle et al., 1999; Wechsler, 2009). For these tasks, participants are required to store the phonological information in short-term memory and, as needed, rehearse it verbatim to ensure accurate responses.

Unlike short-term memory, which does not require the manipulation of information, working memory requires participants to actively process the information. Working memory can be assessed with more complex span tasks, as they require active manipulation of the contents. For example, *backwards digit span* tasks require participants to accurately repeat a series of digits in the reverse order. Another type of working memory task is the *Operation span* task (OSpan; Unsworth et al., 2005), which requires participants to complete an additional, unrelated intervening task between each to-be-remembered item. During the OSpan task, participants must retain and verbally recall a series of letters while solving interspersed math equations (e.g., participant sees a letter, then solves a math question, and then sees another letter, then solves another math question, and so on). At the end of a 2- to 7-item trial set, for example, participants are asked to recall the letters aloud in the order of occurrence. The dual nature of complex span tasks, in comparison to simple span tasks, provides a distinction between storage and rehearsal in working memory and the central executive processing required for attentional control.

A principal component within Baddeley’s model is the central executive (i.e., attentional control system), which directs and allocates attentional resources. However, there has been considerable debate in the literature about the concept of and/or processes involved in attention (Cowan et al., 2005; Chun et al., 2011; Kaldas, 2022). Some researchers have explained attention succinctly as “everybody knows what attention is” (James, 1890). In contrast, others have suggested that “no one knows, or can ever know, exactly what attention is” (Hommel et al., 2019, p. 2299), given its near-ubiquitous impact in any behavioral task. Although not a core component of executive functions (Diamond, 2013), attention is generally considered the foundational, multifaceted cognitive construct that supports the operations of the executive function components when engaged in a purposeful behavior (Baddeley, 2002).

According to some researchers, there are three subcomponents of attention: alerting (also known as sustained, vigilance, or alertness) attention, which is considered the fundamental form of attention and involves being ready to respond to a stimulus about to appear; orienting (also known as selection or scanning) attention, which is related to using a cue to locate a target stimulus, and executive (also known as focused, selective, supervisory, or conflict resolution) attention, which is involved in monitoring and resolving conflicts (Fan et al., 2002; Posner & Boies, 1971; Raz & Buhle, 2006).

Regardless of the different classifications and/or terminologies related to attention as an underlying cognitive construct, Doneva (2020) conducted a meta-analytical review of attentional skills in AWS and AWNS. The results showed that, when compared to AWNS, AWS were less effective and/or less efficient on behavioral attentional tasks that involved dual tasks (12 studies), selective (also known as focused or executive) attention (9 studies), and general attention (21 studies), suggesting that attention may play a role in the persistence of stuttering in adults. In light of these results, the current study focuses on the primary components of executive function, while acknowledging that these constructs (attention and the core executive function components) are highly interrelated and not entirely independent.

### Inhibition

The second core executive function skill is inhibition – the ability to intentionally resist the tendency to execute a dominant or automatic response (Miyake et al., 2000; see also Banich & Depue, 2015; Hall & Fong, 2015). Researchers have classified inhibition based on different theoretical taxonomies or empirical functions (e.g., Friedman & Miyake, 2004; Howard et al., 2014; Nigg, 2000).

Nigg (2000) classified inhibition (also termed executive inhibition) into four distinct conceptual subcategories: (a) interference control, (b) behavioral/motor inhibition, (c) cognitive inhibition, and (d) oculomotor inhibition. Some of these subcategories of inhibition have been theorized to influence stuttered speech at different stages, particularly interference control (e.g., the speaker has difficulty disengaging with inaccurate speech motor plans) and behavioral/motor inhibition (e.g., the speaker has difficulty suppressing an inaccurate speech plan).

According to Nigg’s classification, *Interference control* refers to the ability to maintain one’s response in the presence of a competing, distracting, or interfering stimulus (Dempster, 1993)^1^. There are conceptually different tasks that involve interference control. For example, the Eriksen *flanker task* (Eriksen & Eriksen, 1974) requires participants to press a left/right arrow button in response to a centered targeted stimulus (arrow, >) flanked by other non-target stimuli (arrows; < >) in the same direction (congruent condition; >>> > >>>) or opposite direction (incongruent condition; <<< > <<<).

Response time is the difference in the mean response time between the congruent and incongruent trials (Eriksen, 1995), and response latency is thought to reflect the participants’ ability to resist irrelevant stimuli to achieve a specific goal.

*Behavioral/motor inhibition* refers to a conscious suppression of a dominant behavioral response or movement during a task, such as the Go-NoGo task or Stop-signal tasks.^2^ During a s*top-signal* task (SST), participants press a corresponding button (e.g., the right button for X and the left button for O) as fast as possible, but withhold response when an auditory/visual signal follows the target (Logan, 1994; Verbruggen & Logan, 2008). The participants’ stop-signal reaction time (SSRT) is thought to reflect the ability to suppress a dominant or automatic response after it has been initiated.

*Cognitive inhibition* refers to the ability to effortfully resist previously useful information, thoughts, or directives from intruding into working memory (Dempster, 1993; Harnishfeger, 1995). For example, during a *cued recall* task (Tehan & Humphreys, 1995; Tolan & Tehan, 1999), participants are required to quickly and accurately retrieve a target word from working memory from the most recent block of stimuli while ignoring phonologically-or semantically-related information from preceding trials.

*Oculomotor inhibition* refers to the active suppression of an oculomotor response to gaze towards visual cues flashing in a different location within their visual field (Everling & Fischer, 1998; Hallett, 1978; Hutton & Ettinger, 2006; Magnusdottir et al., 2019).

### Cognitive Flexibility

Cognitive flexibility, the third core executive function component, refers to the ability to adapt or switch one’s responses based on updated rules, strategies, and perspectives (Miyake et al., 2000). Cognitive flexibility typically emerges gradually during the preschool years between 3 and 5 years old (Cragg & Chevalier, 2012; Doebbels & Zelazo, 2015; Ferguson et al., 2021), and its development is considered to be contingent on the development of working memory and inhibition skills (Davidson et al., 2006; Garon et al., 2008). Common behavioral measures of cognitive flexibility are the *Trail Making Test* (Bowie & Harvey, 2006) in which participants connect a sequence of numbers in ascending order (Part A) and subsequently connect alternating numbers and letters (Part B), and the *Wisconsin Card Sorting Task* (Grant & Berg, 1948), wherein participants sort cards based on predetermined set of rules (e.g., sort by color), and then switch to a new rule (e.g., sort by shape). The type and frequency of errors are thought to reflect participants’ ability to adapt or modify their responses after new information is presented and/or errors have occurred.

Cognitive flexibility as a subcomponent of executive function is distinct from, yet interrelated with, working memory and inhibition. In the literature, various terms have been used to refer to cognitive flexibility, including set shifting, task shifting/switching, cognitive adaptability, and mental/representational flexibility (Ionescu, 2012). Whereas working memory can be viewed as the “mental workspace” (i.e., activates, maintains, and processes the information necessary for a goal-directed behavior) and inhibition can be conceptualized as the “mental brake” (i.e., stop or cancel a failed and/or irrelevant goal-directed behavior), cognitive flexibility can be conceptualized as the “mental pivot” – that is, the ability to quickly revise an original goal in response to changes in external or internal environment. In this way, cognitive flexibility relies on working memory (e.g., the storage and processing of stimulus-response patterns) and inhibition (e.g., the suppression of the original information or resultant behavior; Garon et al., 2008). That said, cognitive flexibility has been linked to numerous aspects of mental health (e.g., emotional regulation [Baas et al., 2008; Kashdan & Rottenberg, 2008], resilience [Nakhostin-Khayyat et al., 2024], anxiety/depression mitigation [Johnco et al., 2013; Wang & Lui, 2017]), academic and cognitive performance (e.g., problem solving [Hopper et al., 2020], literacy [Magalhães et al., 2020; Johann et al., 2020], divergent thinking [Baas et al., 2008]), and social functioning (e.g., empathy [Yan et al., 2020]; domestic and marital well-being [Howlett et al., 2022; Koesten et al., 2009]).

### Executive Function Skills in Adults who Stutter

Speech-language planning and production is a complex and dynamic task that involves the efficient, effective, and timely coordination and monitoring of multiple interacting processes (e.g., lexical retrieval, morphological encoding, phonological encoding, monitoring; Dell et al., 2014; Levelt, 1989; Nozari & Novick, 2017). Considering the dynamic and attention-demanding nature of speech-language production, fluent speech production seems to draw upon speakers’ domain-general executive function skills (e.g., Berninger et al., 2017; Shao et al., 2012; Sikora et al., 2016; Ye & Zhou, 2009).

Furthermore, successful conversational interactions depend on the communication partners’ ability to inhibit or suppress internal competing linguistic messages or thoughts (e.g., “I wonder what they think of me when I stutter”) and/or external distractions (e.g., listener reactions, time pressure). For AWS, weaknesses in executive function skills may contribute to an increased cognitive load during speech planning, resulting in greater difficulty sustaining fluent speech.

Furthermore, as previously mentioned, executive function skills are closely associated with psychosocial functioning and quality of life. In the field of stuttering, researchers have reported that AWS often experience psychosocial stressors, reduced socioemotional functioning, and reduced quality of life due to their stuttering (e.g., Blumgart et al., 2010; Boyle, 2015; Craig et al., 2009; Craig & Tran, 2006). Therefore, given the relationship between executive function skills, psychosocial functioning, and quality of life in adults, it is important to examine the executive function skills of AWS. In the stuttering literature, some, but not all, studies conducted with AWS and AWNS suggest that AWS may have less robust executive function skills when compared to AWNS. In the case of working memory, there is evidence that AWS are less accurate during tasks of nonword repetition compared to AWNS (e.g., Byrd et al., 2012; Coalson & Byrd, 2017), suggesting difficulties in phonological working memory; however, other studies have reported no between-talker group differences using similar tasks (e.g., Brown, 2015; Sasisekaran, 2013; Smith et al., 2010). For example, Byrd et al. (2012) examined short-term verbal memory in 14 AWS and 14 AWNS using a modified version of Dollaghan and Campbell’s (1998) nonword repetition task at 4- and 7-syllable lengths, but found between-group differences only at the 7-syllable length level, suggesting that AWS may exhibit weaknesses in phonological working memory. On the other hand, Coalson and Byrd (2017) found between-group differences (26 AWS, 26 AWNS) in nonword recall accuracy for stimuli as short as 2 syllables in length. A similar pattern is reflected in findings across studies that examined inhibition, in which some studies have reported that AWS have weaknesses in behavioral inhibition when compared to AWNS (e.g., Markett et al., 2016), while others have failed to find any significant between-group differences (Bakhtair & Eggers, 2023). Yet still, others also failed to find any differences in oculomotor inhibition between the two groups (e.g., Gkalitsiou et al., 2020). To illustrate, Markett et al. (2016) examined behavioral inhibition between 28 AWS and 28 AWNS using a manual-response stop-signal task and found statistically significant differences between the two groups in response time, but not in accuracy, suggesting that AWS may exhibit weaknesses in behavioral inhibition. Conversely, Bakhtiar and Eggers (2023) did not find any significant differences between 13 AWS and 14 AWNS in the accuracy or reaction time on a verbal stop-signal task. Similarly, Gkalitsiou et al. (2020) did not observe significant differences between 17 AWS and 17 AWNS in the accuracy or reaction time on an antisaccade task, indicating comparable inhibition skills between the two groups.

One possible reason for the discrepancies across individual studies may be variations in task selection and sample size, which can make it more challenging to detect between-group differences. However, by synthesizing and analyzing conceptually similar tasks across individual studies, we can increase the sample size and increase the likelihood of detecting between-group differences, if they do exist. It is possible that between-group differences in executive function skills exist, but to date, individual studies may have been insufficiently powered to yield consistent patterns in findings. It is also equally possible that AWS exhibit no significant differences in executive function skills, or specific subdomains of executive function, compared to AWNS. Thus, the purpose of the present study is to consolidate and summarize the extant quantitative data across conceptually similar tasks in the literature on executive function skills in AWS and AWNS, thereby enhancing the power to detect subtle differences between the two groups to clarify the role of executive function in AWS. More specifically, the quantitative data from empirical studies of working memory, inhibition, and cognitive flexibility between AWS and AWNS will be identified using a search strategy, and the quantitative data related to the conceptually related tasks will be extracted, synthesized, and analyzed. Based on the findings from the Ofoe et al. (2018) study and the evidence that executive function skills continue to develop into adulthood (Ferguson et al., 2021), it is expected that, as a group, AWS will exhibit weaknesses in executive function skills compared to AWNS. Specifically, the following three research questions were examined:

1. Do AWS differ from AWNS on behavioral measures of working memory?
2. Do AWS differ from AWNS on behavioral measures of inhibition?
3. Do AWS differ from AWNS on behavioral measures of cognitive flexibility?

## Method

### Inclusion Criteria

This study was not subject to an Institutional Review Board approval process. To be included in the meta-analysis, a study had to (a) include adults at least 18 years old who do and do not stutter, with the two talker groups not being matched or equated on one or more measures of executive function (talker group classification criteria were free to vary across studies, see Appendix A for summary of group classification criteria); (b) report methods and results in English, and (c) provide quantitative information (e.g., mean and standard deviation) amenable to effect size calculation for at least one core component of executive function (e.g., working memory, cognitive flexibility, and inhibition). Studies that included working memory tasks as secondary tasks to a primary monitoring task were excluded (e.g., phoneme monitoring tasks). In cases where two or more studies reported on overlapping samples (e.g., Eichorn et al., 2016, 2023), only the study with the bigger sample size (e.g., Eichorn et al., 2016) and/or the one with more measures/outcomes was included, to ensure independence of effect sizes. To mitigate publication bias, increase the sample size to improve statistical power, and strengthen the generalizability of the findings, both published (e.g., peer-reviewed journal articles) and unpublished studies (e.g., theses, dissertations) were included.

### Data Sources and Study Selection

The literature search was conducted in accordance with the PRISMA guidelines by the third author, a librarian with experience in conducting systematic reviews, in consultation with the first two authors, primarily using the MEDLINE® database searched via the Ovid interface and the ProQuest Dissertations & Theses Global database. A preliminary MEDLINE search was completed in February 2022, and based on team discussions, further iterations of the searches were conducted in May 2022 and a year later, in June 2023, to capture any newly published studies prior to finalizing the review. Final search strategies reflecting controlled vocabulary terms, keywords, and the search functionality employed (e.g., truncation, field-specific searching, proximity operators) are provided in Appendix B.

The MEDLINE search strategy combined controlled vocabulary (e.g., **“Stuttering/”, “Executive Function/”**) with keyword terms and Boolean logic (e.g., **“stutter$.mp.” OR “stammer$.mp.”**, and **“exec$ function$” OR “memor$ adj2 working$”**) to capture a broad range of relevant literature on stuttering and executive functioning. Search functionalities such as truncation (e.g., **stutter$**, **inhibit$**), proximity operators (e.g., **adj2**), and field-specific searching were employed. A detailed account of the complete search strategy is available in **Appendix B** to ensure transparency and reproducibility.

To ensure comprehensive coverage of dissertations and theses, additional searches were completed in May 2022 by the first two authors using the following sources: Networked Digital Library of Theses and Dissertations (https://ndltd.org/); Open Access Theses and Dissertations (https://oatd.org/); and the Louisiana State University Scholarly Repository (https://repository.lsu.edu/about.html). Limits to English or other languages, publication date, or publication type were not utilized in any of the searches completed for this project.

All identified studies were initially screened based on their title and abstract in Covidence®, a web-based program for systematically managing studies in a meta-analytic review. The first and second authors screened 58% and 42% of the identified studies, respectively, using the outlined criteria. For reliability purposes, each author also screened a subset (20% of studies) of the other rater’s studies. The interrater agreement was excellent (Kappa = 0.91), and any discrepancies were discussed between the two raters until consensus was reached. Full texts of all studies that passed the first level of screening (N = 98) were then retrieved and were independently reviewed for final inclusion by the first two authors and the fourth author. All cases of disagreement in eligibility assessment were resolved through detailed discussion between all three raters.

### Data Management and Extraction

A summary of the number of studies excluded during the review process is provided in Figure 1. A total of 39 studies qualified for inclusion. Descriptive data (e.g., age range of participants, construct(s) measured, task description) and quantitative data (e.g., sample sizes, means, standard deviations, statistical tests) were extracted from each of the 39 studies that met inclusion criteria, and they were manually coded into a Microsoft Excel spreadsheet. The descriptive data for all included studies can be found in Table 2.

**Figure 1.**
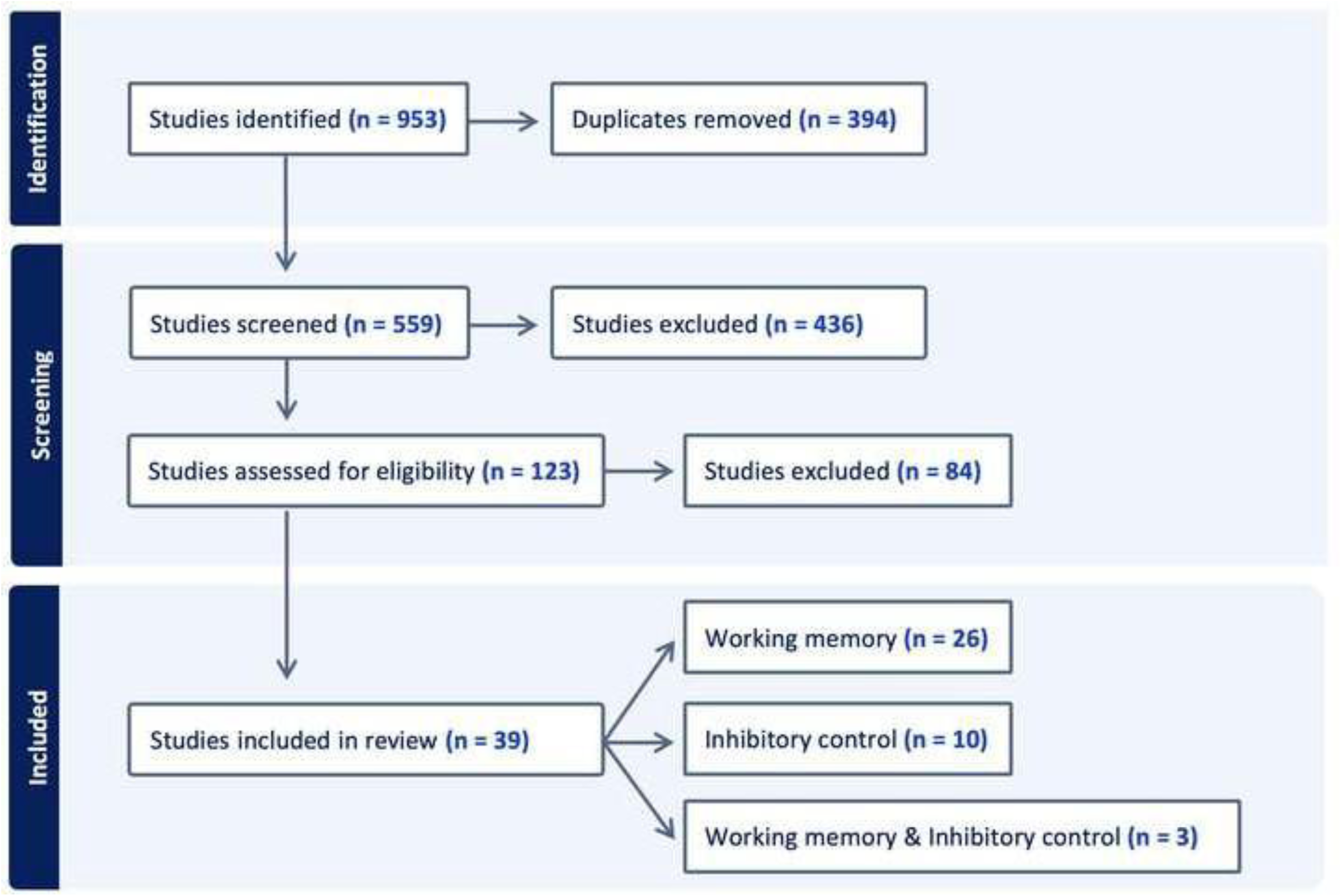
PRISMA flow diagram for systematic review.

### Statistical Analysis

All statistical analyses were performed with the Comprehensive Meta-Analysis (Version 4; Borenstein et al., 2022) software package. Hedges’ *g*, a variation of the standardized mean difference (SMDs) correcting for potential bias due to small sample sizes (Hedges & Olkin, 1985), and 95% confidence intervals (CI) were calculated for each study and construct (see Appendix C for the glossary). When multiple levels of a single measure were extracted from a study (e.g., nonword repetition accuracy for 4-syllable and 7-syllable nonwords), the effect sizes for each level were averaged to yield a single effect size for that study. A random effects model was used in all analyses, weighing the studies based on their sample size and standard error.

The Cochrane *Q* statistic, using a criterion alpha of 0.1, instead of the conventional level of 0.05, and *I^2^* were used to assess heterogeneity across studies (Higgins et al., 2003). Higgins et al. (2003) recommend using a significance level of 0.1 rather than the conventional 0.05, due to the limited power of the Cochrane Q test to detect heterogeneity, especially when few studies are included in a meta-analysis. Publication bias was evaluated through a combination of visual inspection of funnel plots, Duval and Tweedie’s trim-and-fill procedure (Duval & Tweedie, 2000), and Egger’s regression test.

Because funnel plot asymmetry can arise from factors other than publication bias (e.g., heterogeneity), the results were interpreted with attention to the direction of any missing studies. Specifically, we considered whether the location of imputed studies aligned with the direction in which bias would be expected to operate (i.e., AWS performing better than AWNS). Finally, we evaluated the robustness of the results for each construct/measure (e.g., nonword repetition) by conducting the “one-study removed” sensitivity analysis. In this approach, the analysis was repeated multiple times, each time excluding one study from the analysis. This iterative process helps determine if any single study disproportionately influences the overall effect size and, consequently, the conclusions are drawn.

## Results

The descriptive characteristics of studies included in the meta-analysis are presented in Table 1.

**Table 1.**
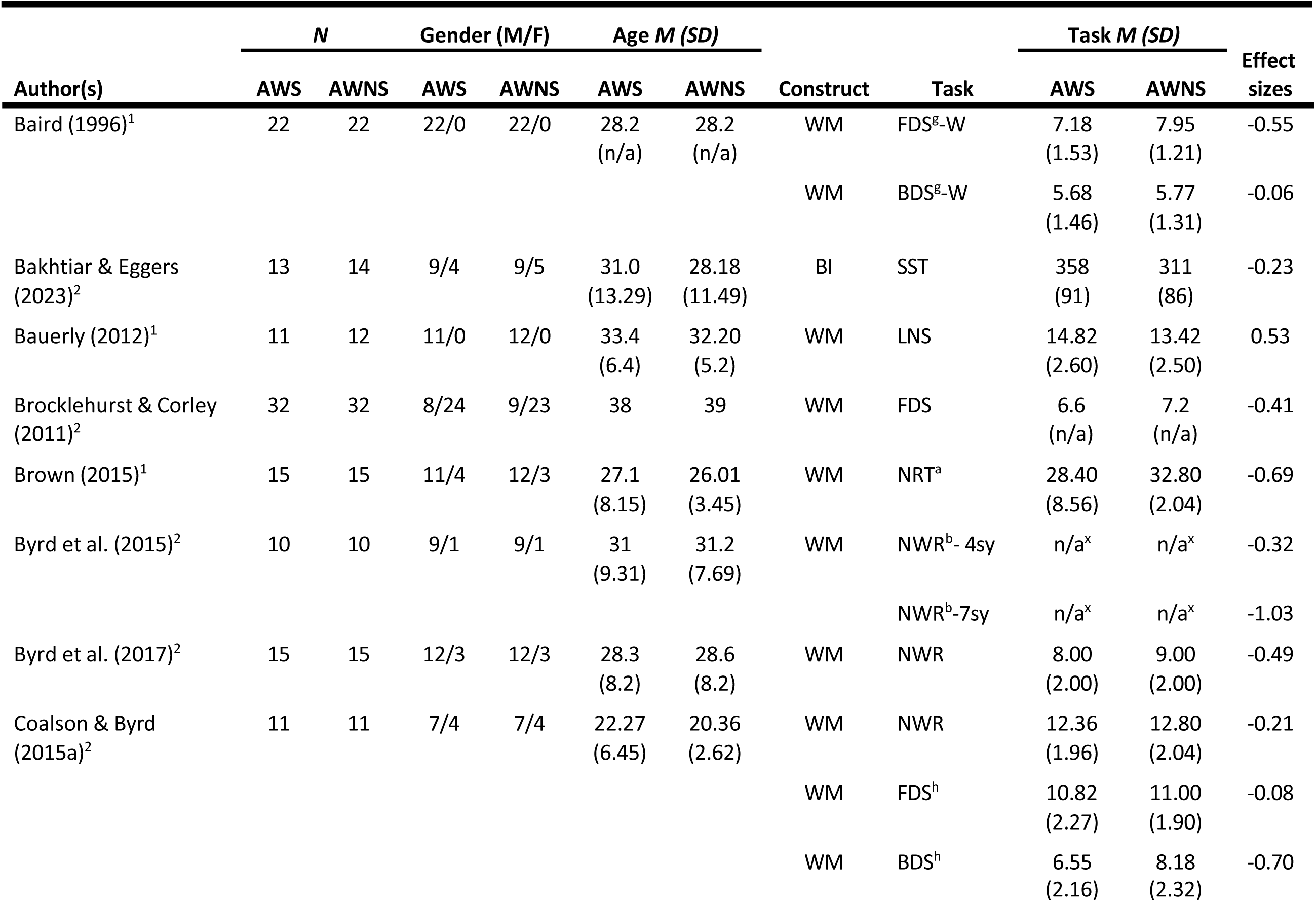
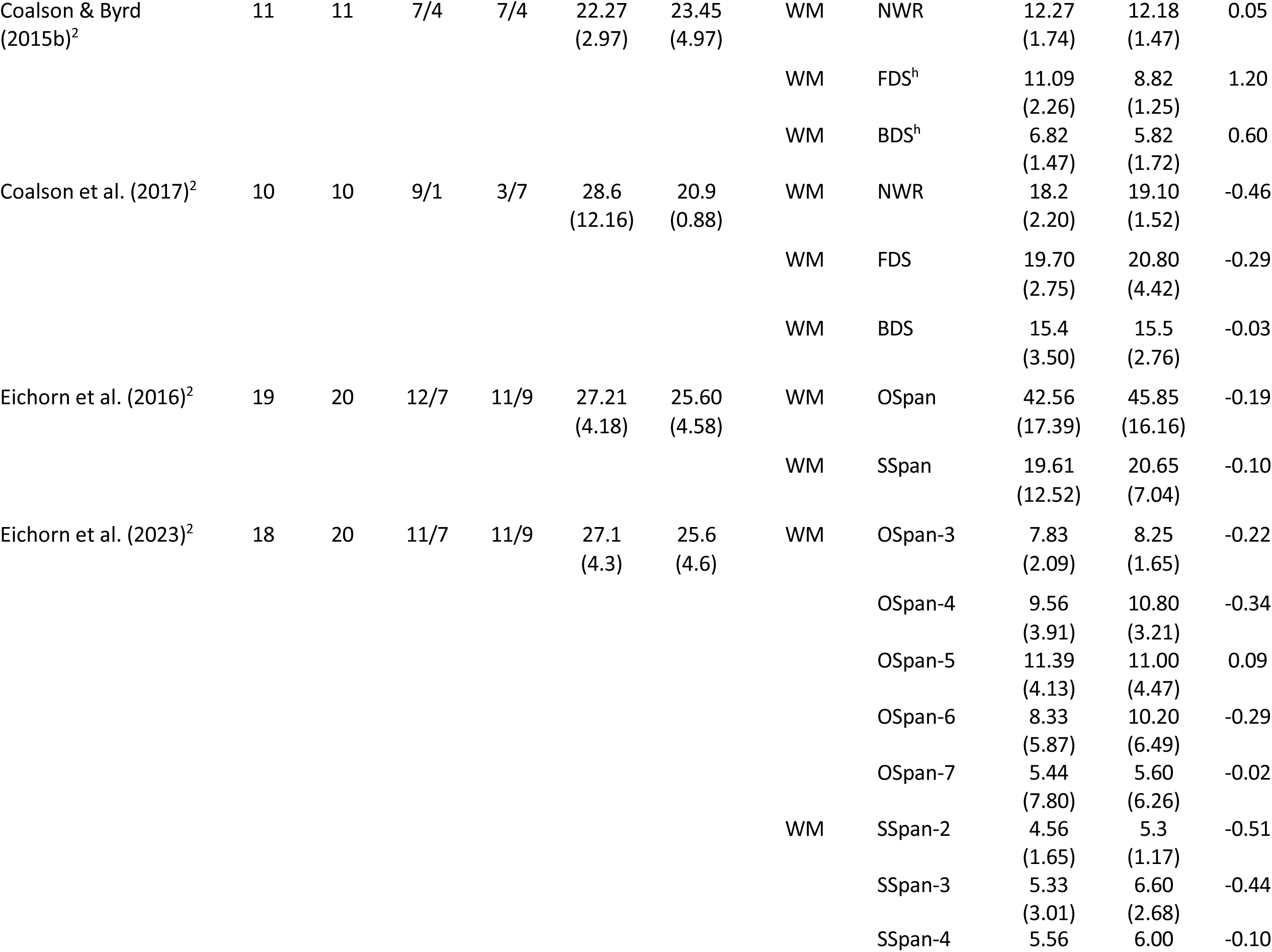
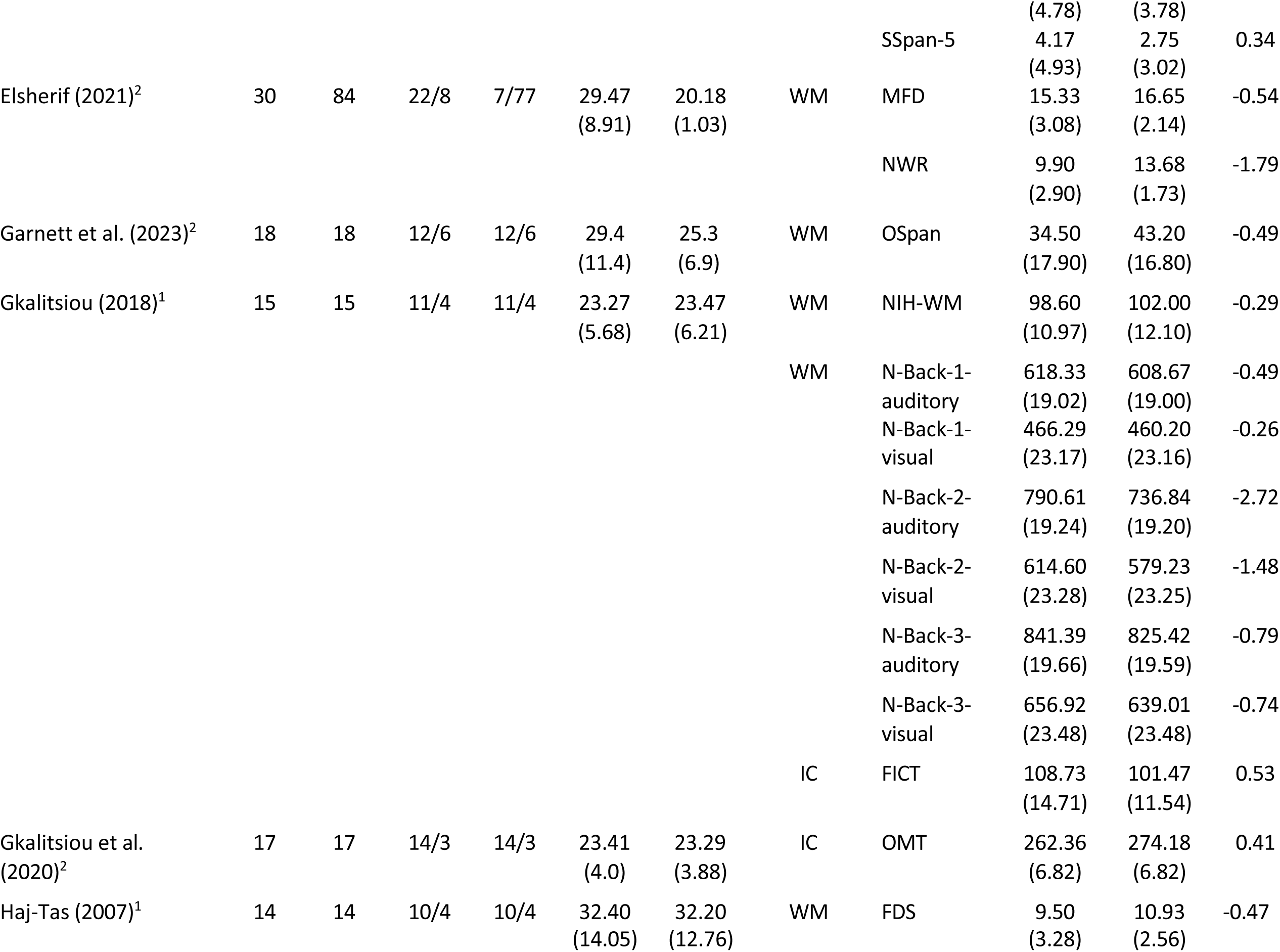
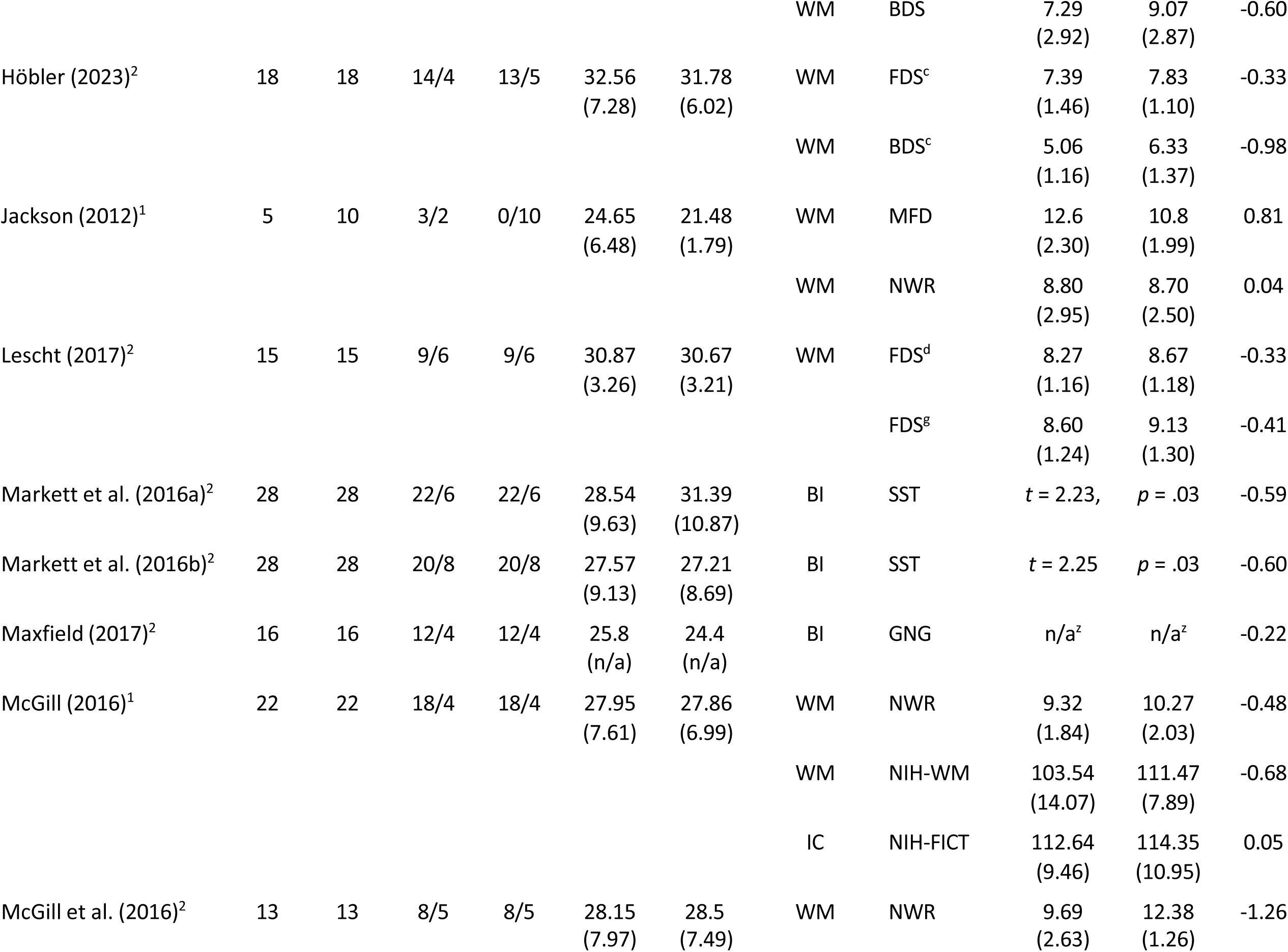
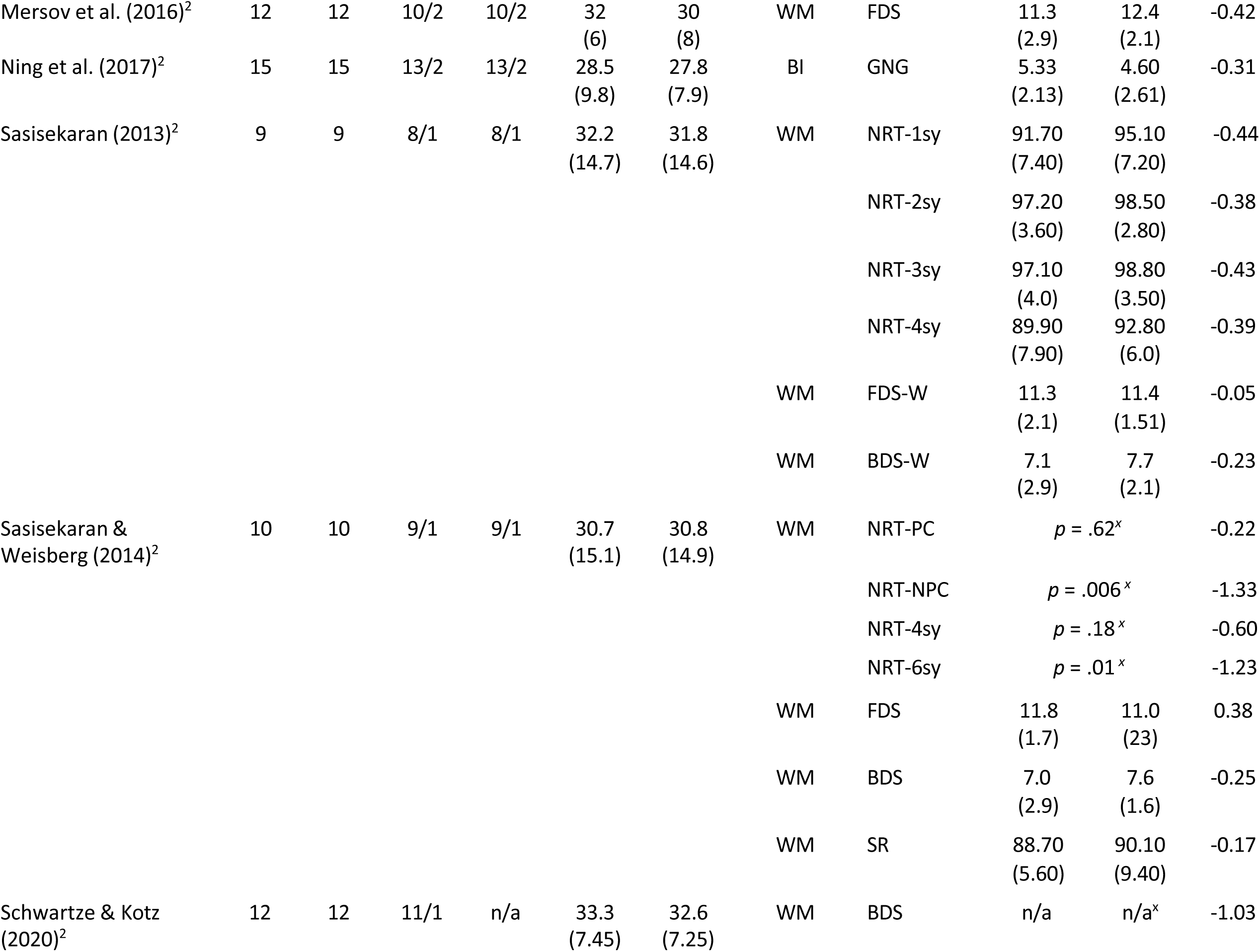
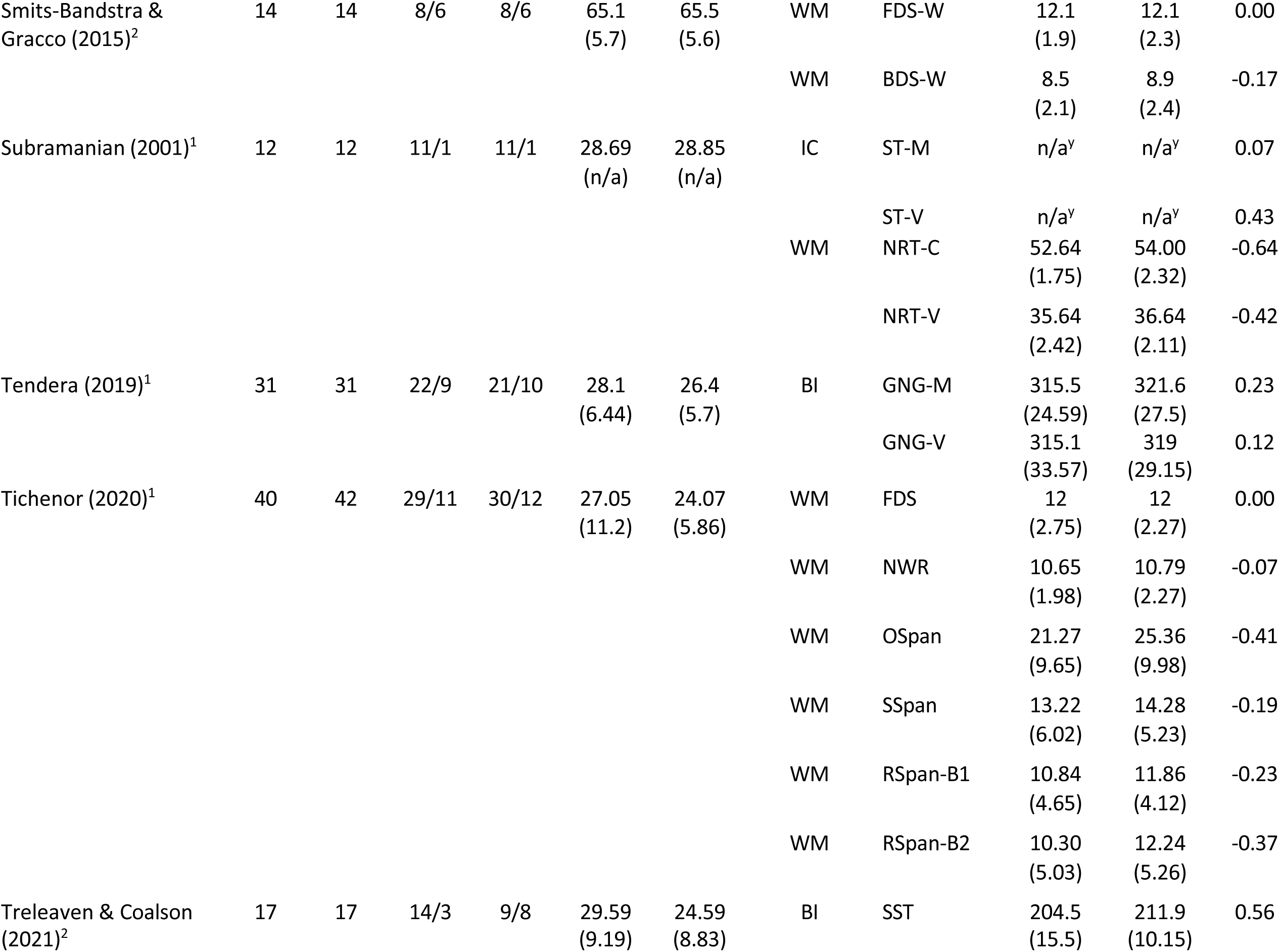
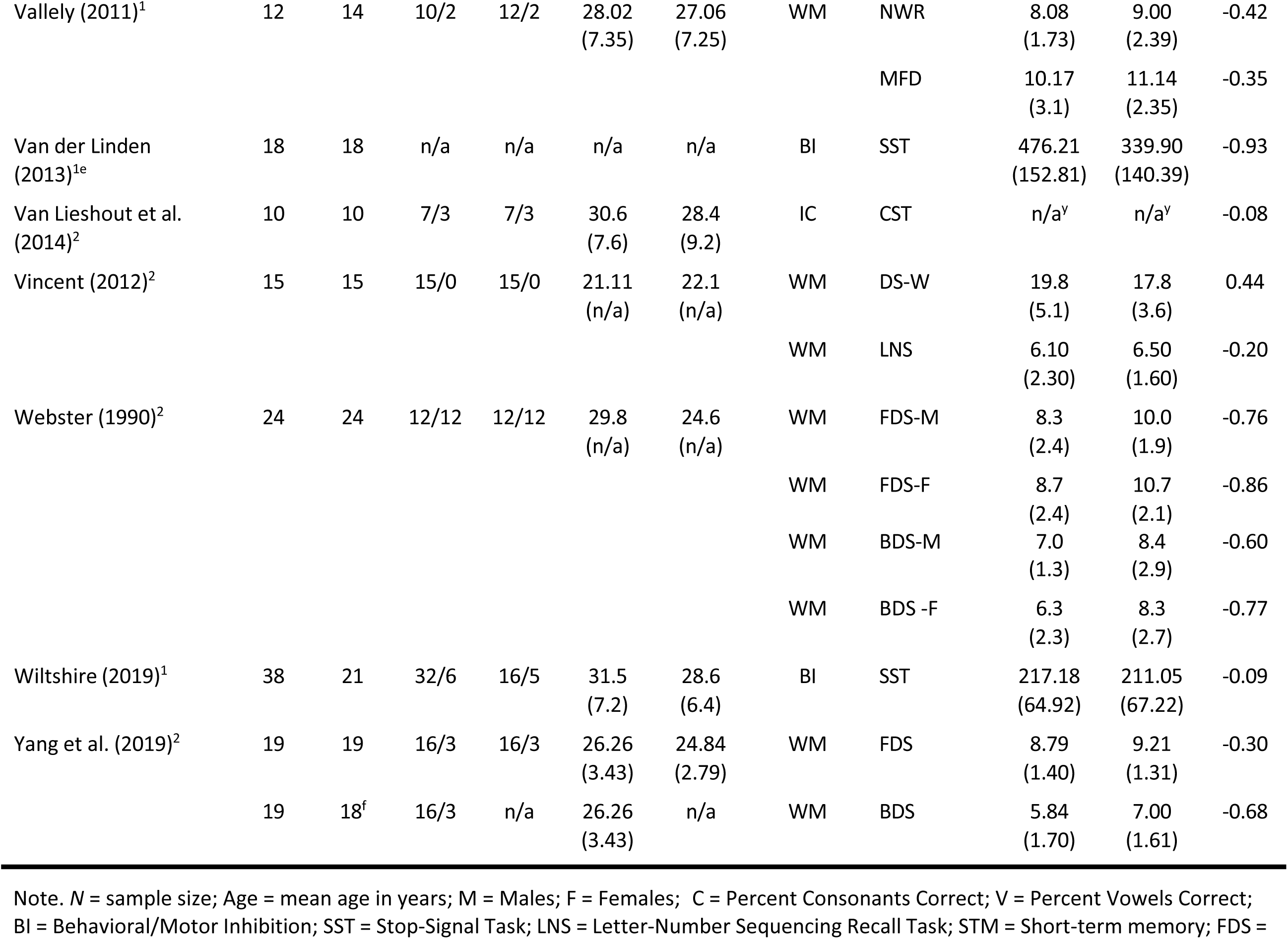
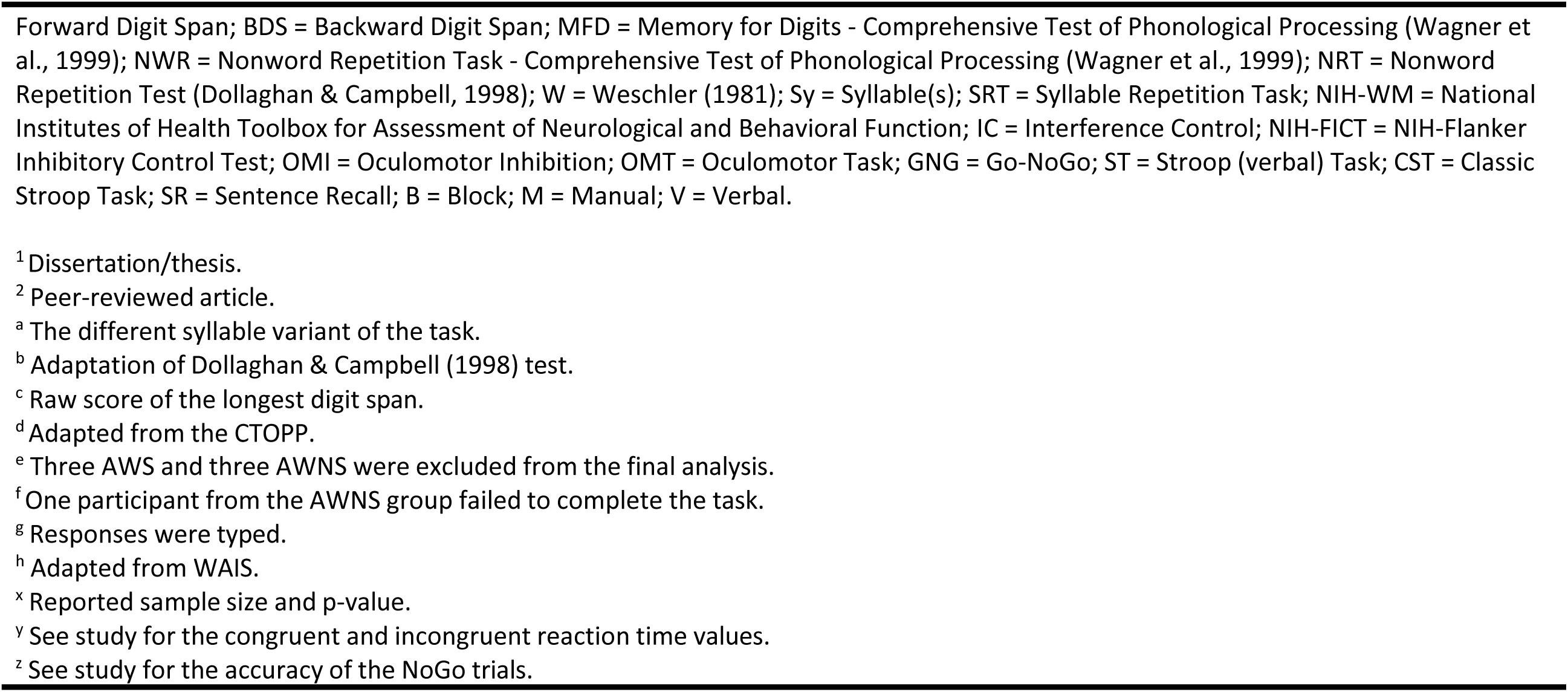
Descriptive Information of Studies that examined Executive Function between Adults Who Do (AWS) and Do Not Stutter (AWNS).

The Coalson and Byrd (2015) study included two independent samples, so it provided two separate effect size estimates for the analyses. Of the 39 studies included in the analysis, 23 (59%) were conducted in the United States, seven (17.9%) in Canada, three (7.7%) in the United Kingdom, three (7.7%) in China, two (5.1%) in Germany, and one (2.6%) in The Netherlands. Twenty-seven (75%) of the 39 studies were reported in peer-reviewed journal articles, nine (23.1%) were doctoral dissertations, and three (7.7%) were Master’s theses. In the majority of the studies (69%), participants were assigned to AWS or AWNS talker groups based on traditional stuttering measures (e.g., percent of stuttering-like disfluencies from conversational or reading samples [Yairi & Ambrose, 2005], stuttering severity measures based on the Stuttering Severity Instrument [Riley, 1994; 2009]; see Appendix A). For the remaining studies, group membership was primarily based on self-identification as an adult who stutters and/or by clinical judgment of a speech-language pathologist. Twenty-four studies reported that participants in the AWS and AWNS groups were equated or matched on education. The remaining studies did not provide any information on the educational level of the two talker groups. Of the 39 included studies, 17 measured participants’ vocabulary skills, of which only two (Haj-Tas, 2007; Sasisekaran & Weisberg, 2014) reported a statistically significant difference between AWS and AWNS, with AWS demonstrating lower vocabulary skills. The remaining 15 studies did not find a statistically significant difference in receptive or expressive vocabulary between AWS and AWNS. Finally, only six studies assessed participants’ nonverbal intelligence, with the overwhelming majority using the *Test of Nonverbal Intelligence–Fourth Edition* (TONI-4; Brown et al., 2010), and one study using the *Raven’s Standard Progressive Matrices* (Raven, 1960). None of the studies reported a statistically significant difference in nonverbal intelligence between AWS and AWNS.

Of the 39 total studies, 26 studies included working memory measures, 10 studies included inhibition measures, and three studies included measures for both. An insufficient number of studies on cognitive flexibility were obtained from the search for any meaningful analysis (*n* = 3; Doneva et al., 2018; Gkalitsiou, 2018; Markett et al., 2016). Therefore, quantitative analysis of studies that examined cognitive flexibility was not conducted.

### Working Memory

#### Nonword Repetition

A total of 14 studies examined verbal short-term memory in 225 AWS and 288 AWNS using nonword repetition tasks. As shown in Figure 2, results of the random effect estimate of the mean effect size revealed a weighted moderate effect size of Hedges’ *g* = -.57, *p* < .001 (SE = 0.14, 95% CI [−0.30, - 0.85]). This mean effect size suggests that AWS, as a group, scored more than half a standard deviation below AWNS on nonword repetition measures.

**Figure 2.**
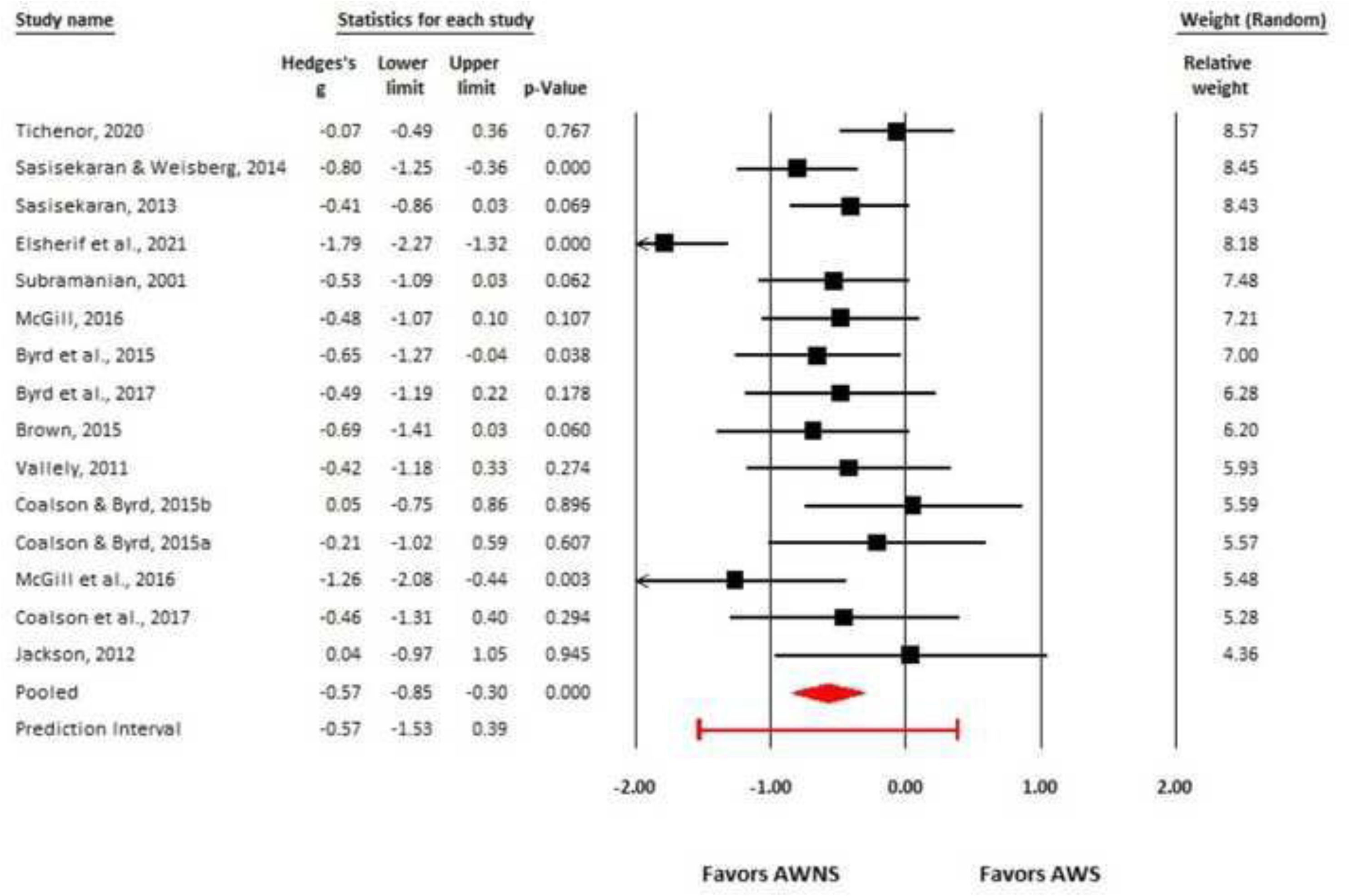
Forest plot for the mean difference in nonword repetition skills between adults who do (AWS) and do not stutter (AWNS). Squares represent effect size per study, with size proportional to its weight in the analysis. Horizontal lines indicate 95% confidence intervals (CIs) per study. The red diamond represents the overall effect size and corresponding 95% CI. The red prediction interval reflects the expected range within which the true effect sizes of future studies fall.

A visual inspection of the funnel plot (see Figure 3) indicated potential evidence of publication bias, and the Duval and Tweedie’s trim and fill method estimated that at least five effect sizes with negative Hedges’ *g*-values would be necessary to make the funnel symmetrical. However, if publication bias were present, it would be more likely to exist against studies showing positive effects (i.e., AWS performing better than AWNS). Because the five missing studies fall in the opposite direction of where bias would be expected, and the Egger’s regression test was nonsignificant (β = 1.22, SE = 1.71, *p* = .24), one could argue that the average effect size estimate for the nonword repetition measure is not affected by publication bias. Other factors (e.g., heterogeneity) could also contribute to funnel asymmetry.

**Figure 3.**
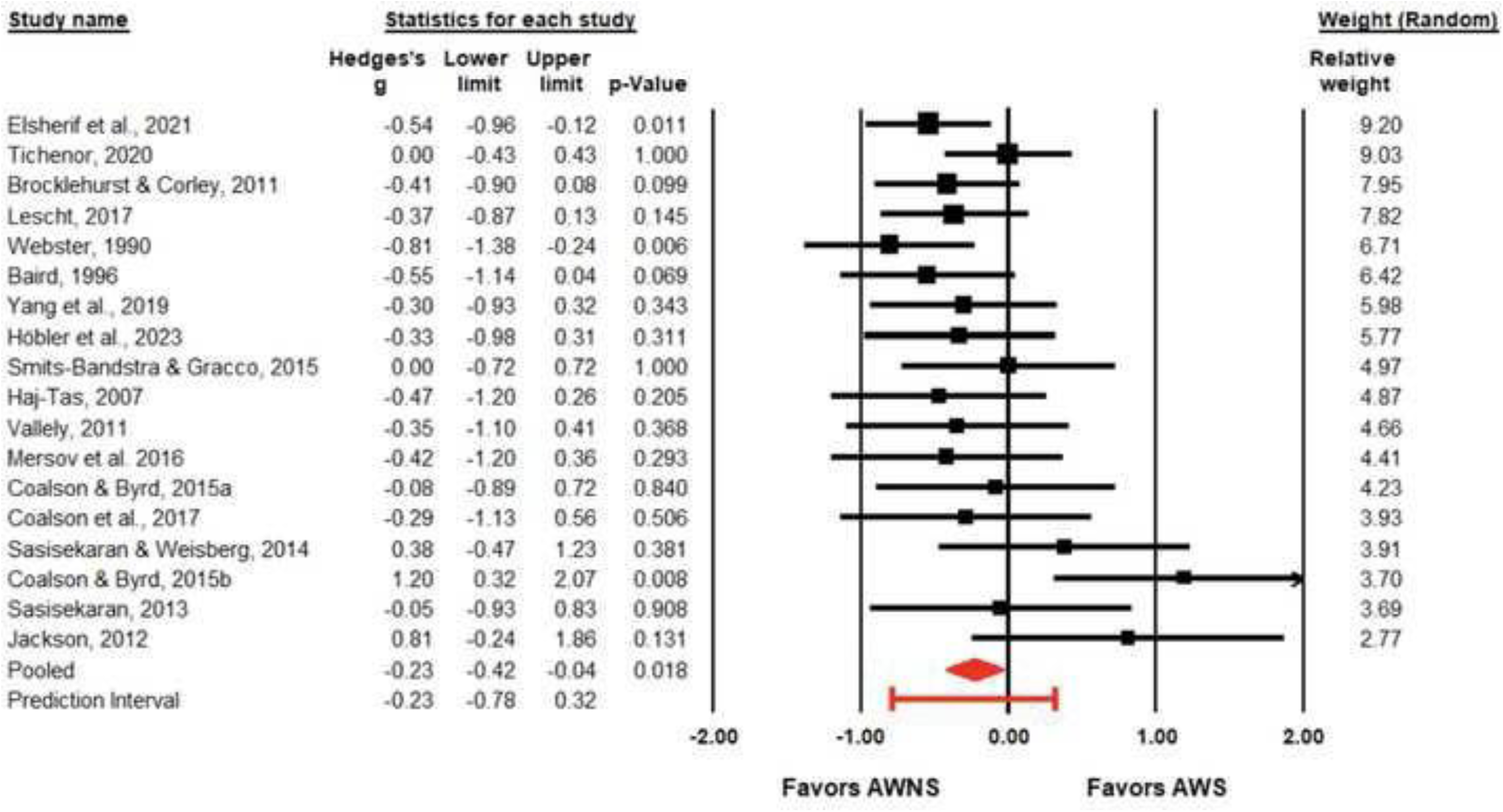
Funnel plot for studies that examined nonword repetition skills of adults who do (AWS) and do not stutter (AWNS). The white circles represent individual studies, and white diamonds represent the pooled estimate. Five imputed studies, based on Duval and Tweedie’s trim and fill method, are shown as filled circles, with filled diamonds reflecting the imputed/“adjusted” Hedges’ *g* estimates.

Analysis of heterogeneity revealed a significant variation in effect sizes across the studies, *Q* = 39.80, df = 14, *p* < .001, suggesting that effect sizes differ more than would be expected by chance alone. The *I^2^* statistic was 64.82%, indicating that approximately 65% of the variance in observed effects reflects variance in true effects rather than sampling error. Based on the prediction interval, shown at the bottom of the forest plot, the true effect size in 95% of all comparable future studies will fall between −1.53 and 0.39. Finally, the direction and magnitude of Hedges’ *g* persisted after sequentially excluding each study, suggesting that the pooled effect size was not greatly affected by any one study.

#### Forward Digit Span

Seventeen studies assessed verbal short-term memory between 308 AWS and 371 AWNS using forward digit span measures. As shown in Figure 4, based on the pooled distribution, results of the random effect estimate of the mean effect size revealed a weighted small effect size of Hedges’ *g* = -.23, *p* = .02 (SE = .09, 95% CI [−0.42, −0.04]). This mean effect size suggests that AWS, as a group, scored about a third of a standard deviation below AWNS on forward digit span measures. A visual inspection of the funnel plot and the Duval and Tweedie’s trim and fill method failed to reveal any evidence of publication bias.

**Figure 4.**
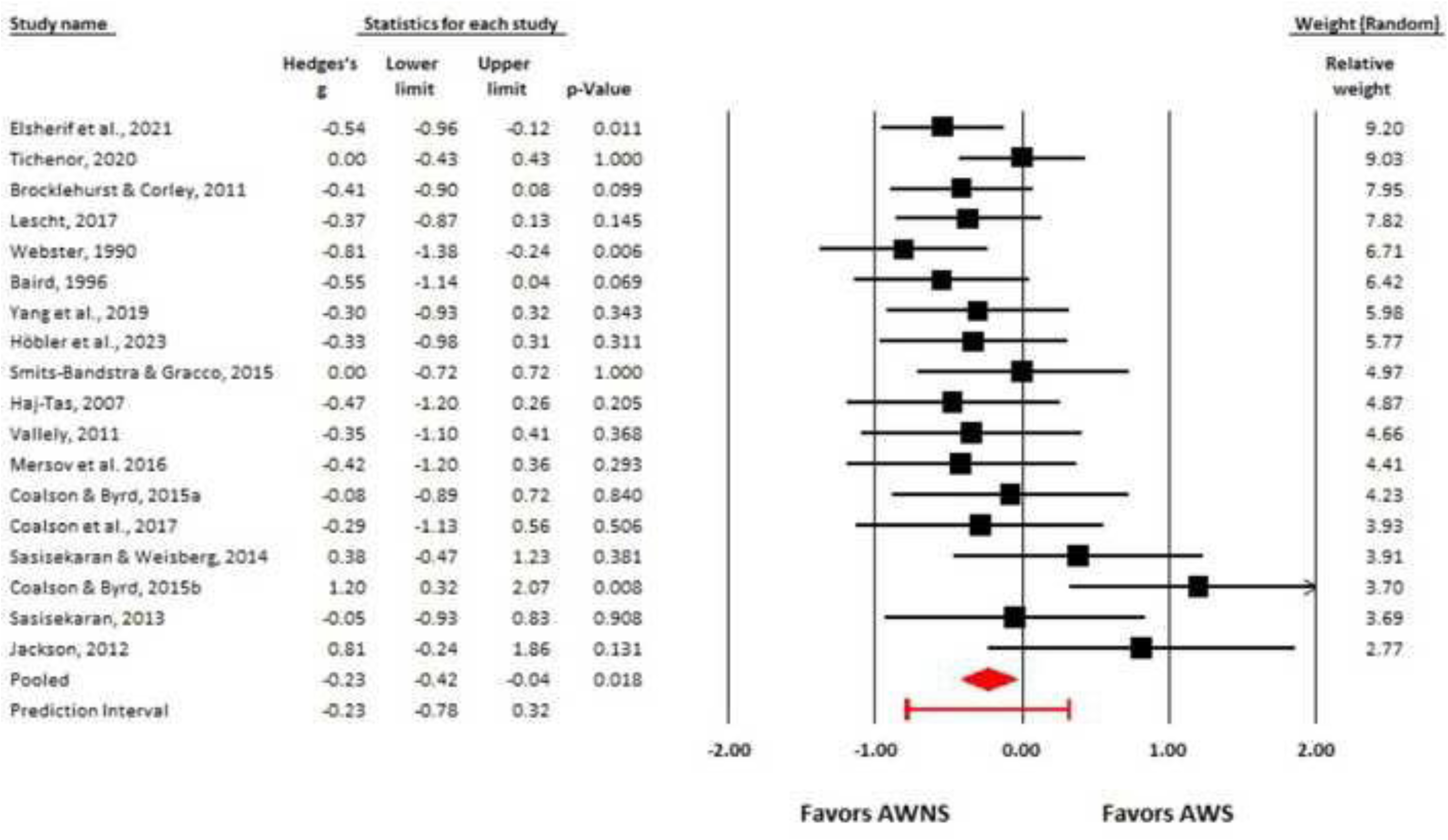
Forest plot for the mean difference in forward digit span skills between adults who do (AWS) and do not stutter (AWNS). Squares represent effect size per study, with size proportional to its weight in the analysis. Horizontal lines indicate 95% confidence intervals (CIs) per study. The red diamond represents the overall effect size and corresponding 95% CI. The red prediction interval reflects the expected range within which the true effect sizes of future studies fall.

Analysis of heterogeneity revealed significant variation in effect sizes across the studies, *Q* = 26.38, df = 17, *p* = .07. Based on the *I^2^* statistic, approximately 35% of the variance in observed effects reflects variance in true effects rather than sampling error. Based on the prediction interval, the true effect size in 95% of all comparable future studies will fall between −0.78 and 0.32. A “one-study removed” sensitivity analysis indicated that the results are robust and stable, as the average effect size remained essentially unchanged with any one study removed.

#### Backward Digit Span

Ten studies assessed verbal working memory between 162 AWS and 161 AWNS using the backward digit span measures. As shown in Figure 5, based on the pooled distribution, results of the random effect estimate of the mean effect size revealed a weighted moderate effect size of Hedges’ *g* = -.38, *p* = .004 (SE = 0.13, 95% CI [−0.64, −0.12]), with AWNS, as a group, outperforming the AWS group. A visual inspection of the funnel plot and the Duval and Tweedie’s trim and fill method did not reveal any evidence of publication bias.

**Figure 5.**
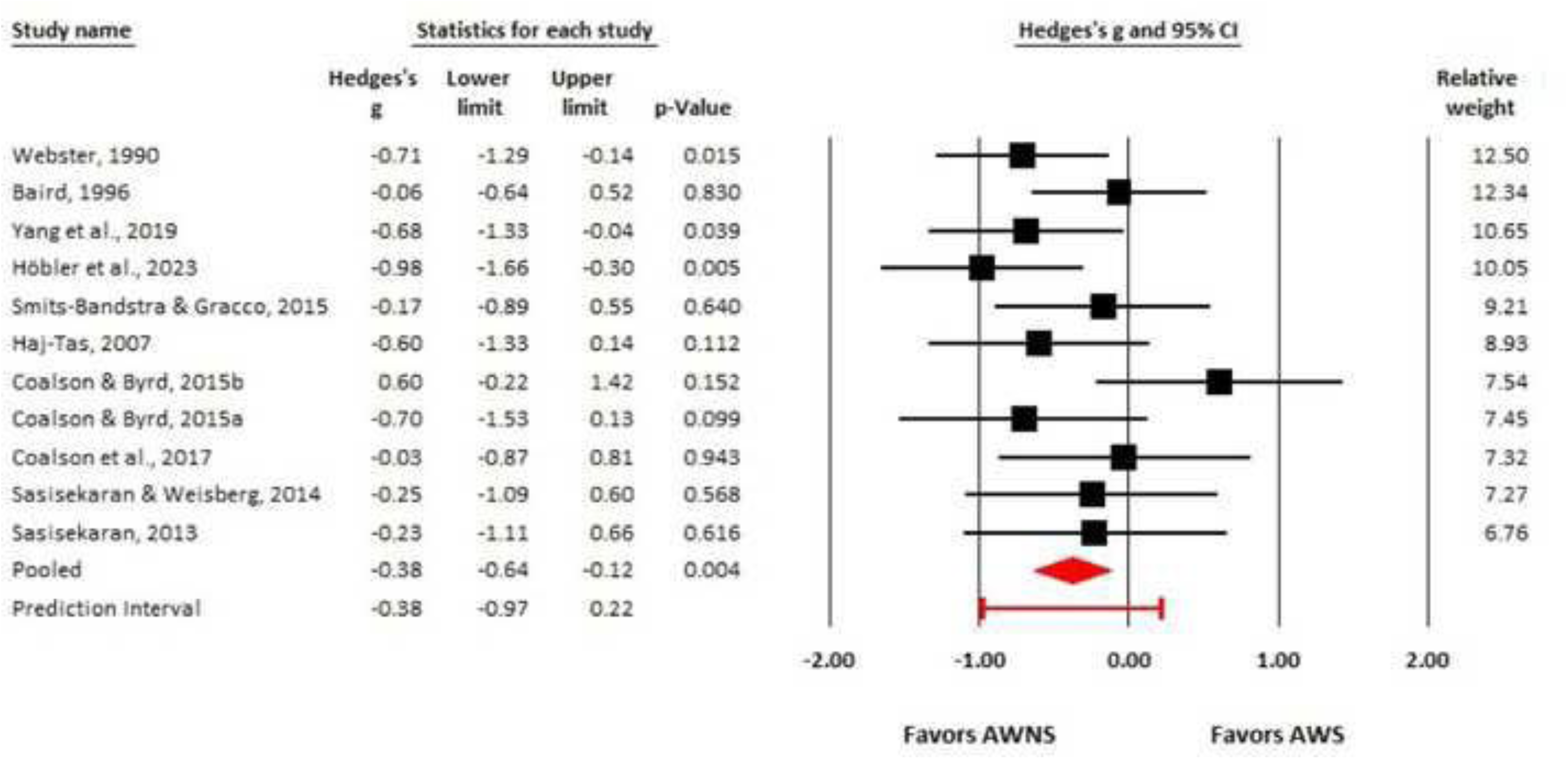
Forest plot for the mean difference in backward digit span skills between adults who do (AWS) and do not stutter (AWNS). Squares represent effect size per study, with size proportional to its weight in the analysis. Horizontal lines indicate 95% confidence intervals (CIs) per study. The red diamond represents the overall effect size and corresponding 95% CI. The red prediction interval reflects the expected range within which the true effect sizes of future studies fall.

**Figure 6.**
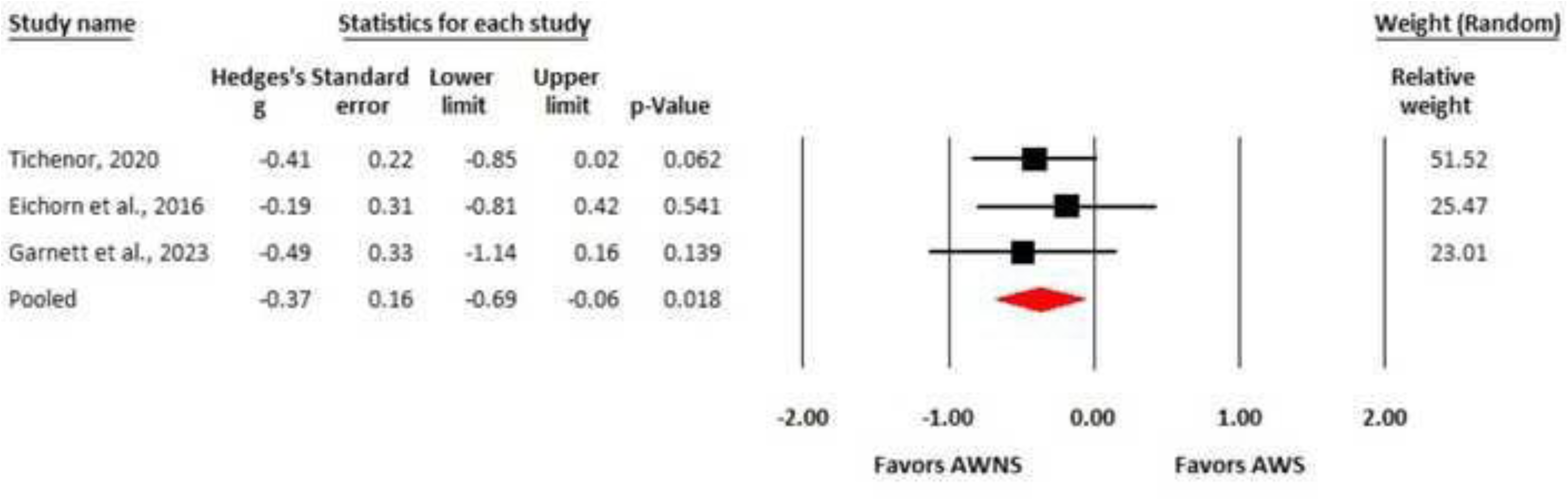
Forest plot for the mean difference in operation span (OSpan) skills between adults who do (AWS) and do not stutter (AWNS). Squares represent effect size per study, with size proportional to its weight in the analysis. Horizontal lines indicate 95% confidence intervals (CIs) per study. The red diamond represents the overall effect size and corresponding 95% CI.

Analysis of heterogeneity did not support a significant variation in effect sizes across the studies, *Q* = 13.83, df = 10, *p* = .18. Based on the *I^2^* statistic, approximately 28% of the variance in observed effects reflects variance in true effects rather than sampling error. Based on the prediction interval, the true effect size in 95% of all comparable future studies will fall between −0.97 and 0.22. Sensitivity analysis with one study removed did not change the average effect size estimate.

#### OSpan

Three studies examined complex verbal working memory skills between 77 AWS and 80 AWNS using the OSpan measures. As shown in Figure 5, based on the pooled distribution, the results of the random effect estimate of the mean effect size revealed a weighted small-moderate effect size of Hedges’ *g* = -.37, *p* = .018 (SE = 0.16, 95% CI [−0.69, −0.06]), with AWS, as a group, scoring lower on OSpan tasks than the AWNS group. A visual inspection of the funnel plot failed to reveal any evidence of publication bias. An analysis of heterogeneity revealed the absence of variability across studies (*I^2^* = 0), *<u>Q</u>* = 0.487, df = 2, *p* = .784. Given the zero between-study variance, a prediction interval was not calculated, and a sensitivity analysis was not performed. However, these results should be interpreted with caution because estimates of heterogeneity based on fewer than ten studies may be unreliable (Borenstein, 2020).

#### Overall Working Memory

Some studies used working memory measures (e.g., list sorting working memory task) that did not fit into the discrete categories of nonword repetition, forward digit span, backward digit span, and OSpan. These measures were combined with all the other measures to generate an overall working memory effect size. In total, 29 studies assessed working memory skills. As shown in Figure 7, the results of the random effect estimate of the overall mean effect size revealed a weighted medium effect size of Hedges’ *g* = -.41 (SE = 0.07, 95% CI [−0.55, −0.27]).

**Figure 7.**
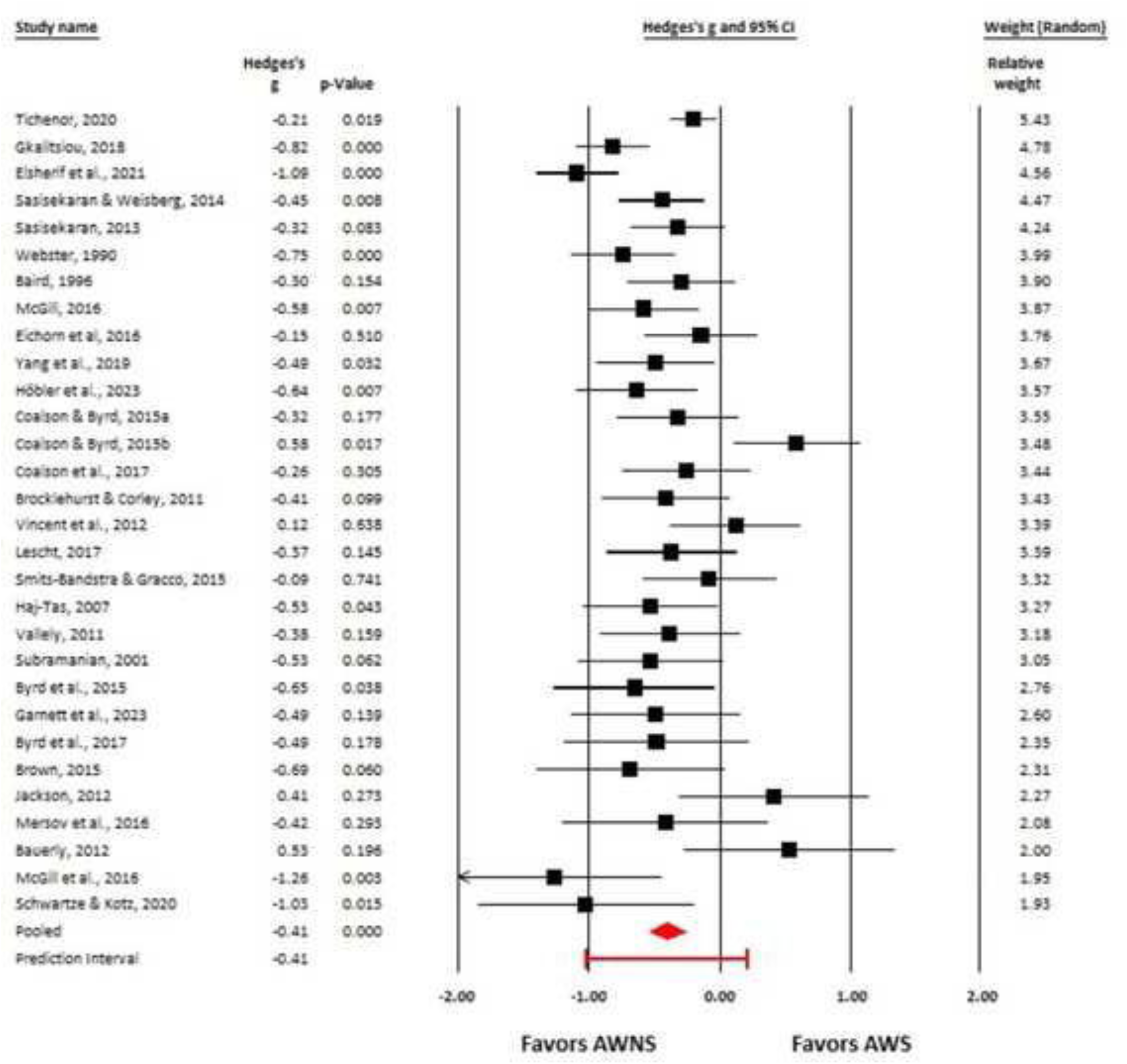
Forest plot for the mean difference in overall working memory skills between adults who do (AWS) and do not stutter (AWNS). Squares represent effect size per study, with size proportional to its weight in the analysis. Horizontal lines indicate 95% confidence intervals (CIs) per study. The red diamond represents the overall effect size and corresponding 95% CI. The red prediction interval reflects the expected range within which the true effect sizes of future studies fall.

A visual inspection of the funnel plot (see Figure 8) indicated potential evidence of publication bias, and the Duval and Tweedie’s trim and fill method estimated that at least seven effect sizes with positive Hedges’ *g*-values would be necessary to make the funnel symmetrical. However, the Egger’s regression test showed a non-significant intercept (β = .06, SE = 0.80, *p* = 0.46), suggesting that the average effect size estimate for studies examining working memory may not necessarily be affected by publication bias.

**Figure 8.**
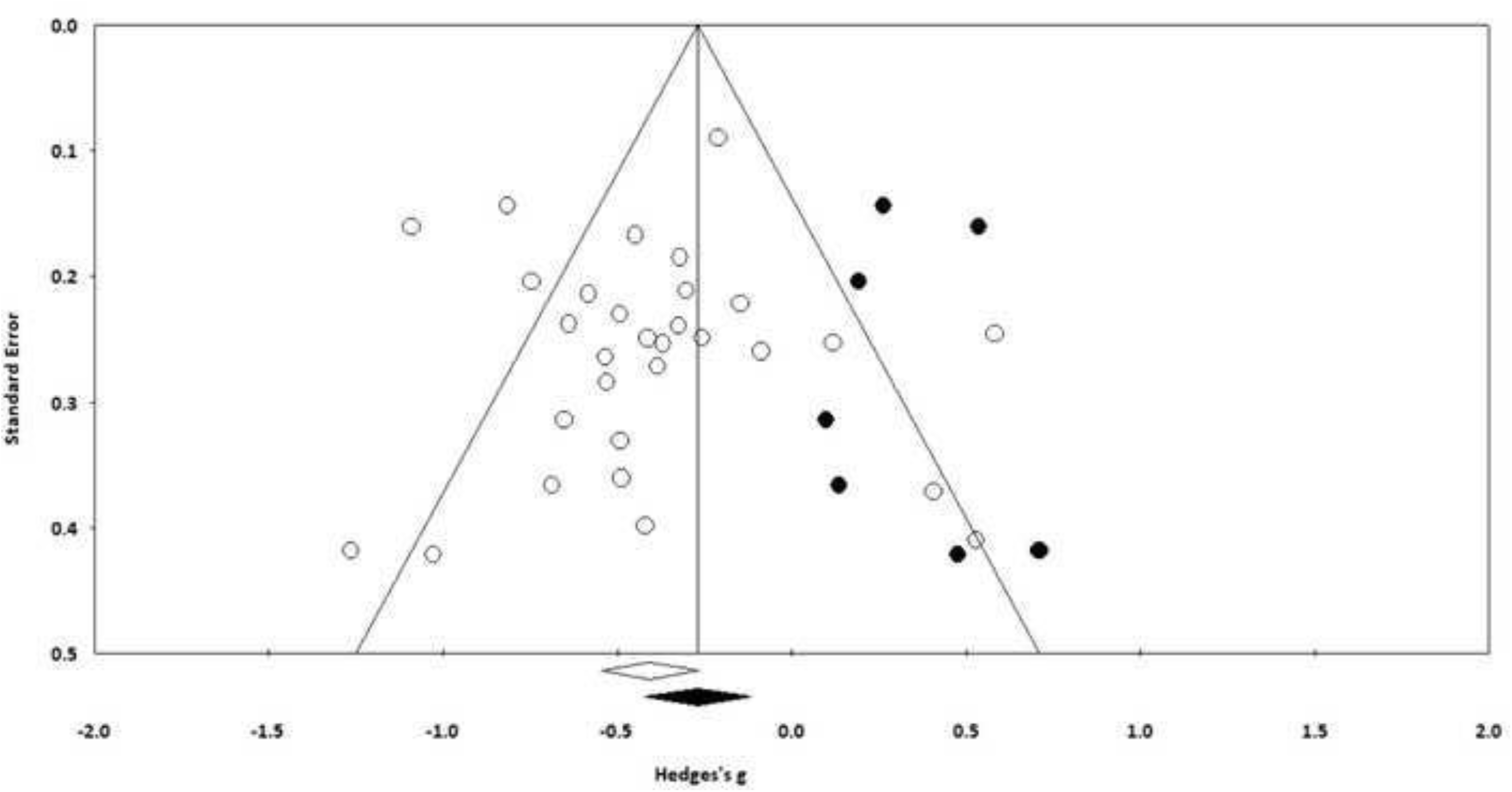
Funnel plot for studies that examined the working memory skills of adults who do (AWS) and do not stutter (AWNS). The white circles represent individual studies, and white diamonds represent the pooled estimate. Five imputed studies, based on Duval and Tweedie’s trim and fill method, are shown as filled circles, with filled diamonds reflecting the imputed/“adjusted” Hedges’ *g* estimates.

Analysis of heterogeneity revealed significant variation in effect sizes across the studies, *Q* = 78.125, df = 30, *p* < .001. Based on the *I^2^* statistic, approximately 63% of the variance in observed effects reflects variance in true effects rather than sampling error. This heterogeneity is not surprising given the different tasks/measures included in the analysis. Based on the prediction interval, the true effect size in 95% of all comparable future studies will fall between −1.02 and 0.21. A “one-study removed” sensitivity analysis indicated that the results are robust and stable, as the average effect size remained essentially unchanged when any one study was removed.

### Inhibition

#### Interference Control

Four studies examined interference control between 59 AWS and 59 AWNS using the Stroop or flanker measures. As shown in Figure 9, results of the random effect estimate of the mean effect size revealed a weighted small and non-significant effect size of Hedges’ *g* = .19 (SE = .15, 95% CI [-.12, .49]). This result suggests that AWS and AWNS do not differ significantly in interference control inhibition. A visual inspection of the funnel plot and the Duval and Tweedie’s trim and fill method did not reveal any evidence of publication bias.

**Figure 9.**
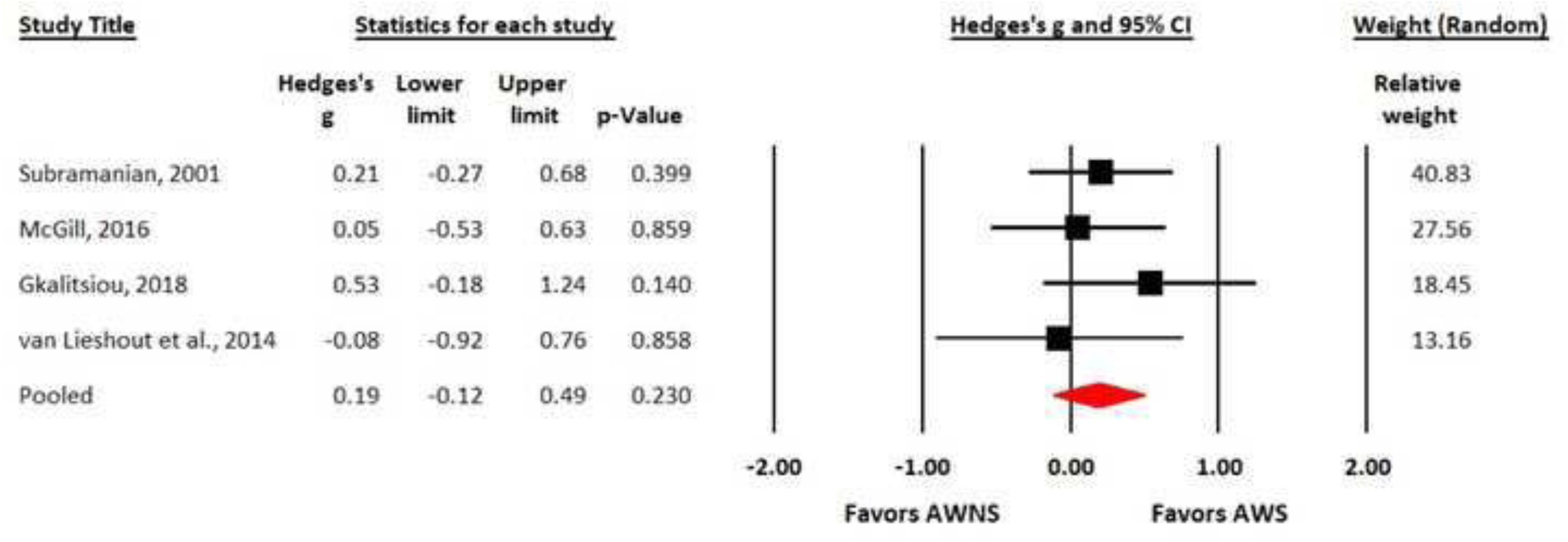
Forest plot for the mean difference in interference inhibition control skills between adults who do (AWS) and do not stutter (AWNS). Squares represent effect size per study, with size proportional to its weight in the analysis. Horizontal lines indicate 95% confidence intervals (CIs) per study. The red diamond represents the overall effect size and corresponding 95% CI.

An analysis of heterogeneity revealed the absence of variability across studies (*I^2^* = 0), *Q* = 1.51, df = 3, *p* = .70. Given the zero between-study variance, a prediction interval was not calculated, and a sensitivity analysis was not performed. However, as previously mentioned, these results need to be interpreted with caution because estimates of heterogeneity based on fewer than ten studies could be unreliable (Borenstein, 2020; Borenstein et al., 2021).

#### Behavioral/Motor Inhibition

Eight studies examined behavioral/motor inhibition of 204 AWS and 188 AWNS using the Go-NoGo and stop-signal measures. The Markett et al. (2016) study included two independent samples, so it provided two separate effect size estimates to the analysis. As shown in Figure 10, based on the pooled distribution, results of the random effect estimate of the mean effect size revealed a non-significant effect size of Hedges’ *g* = -.25 (SE =.14, 95% CI [-.52, .02]). This mean effect size suggests that AWS, as a group, scored similarly to AWNS on behavioral/motor inhibition measures.

**Figure 10.**
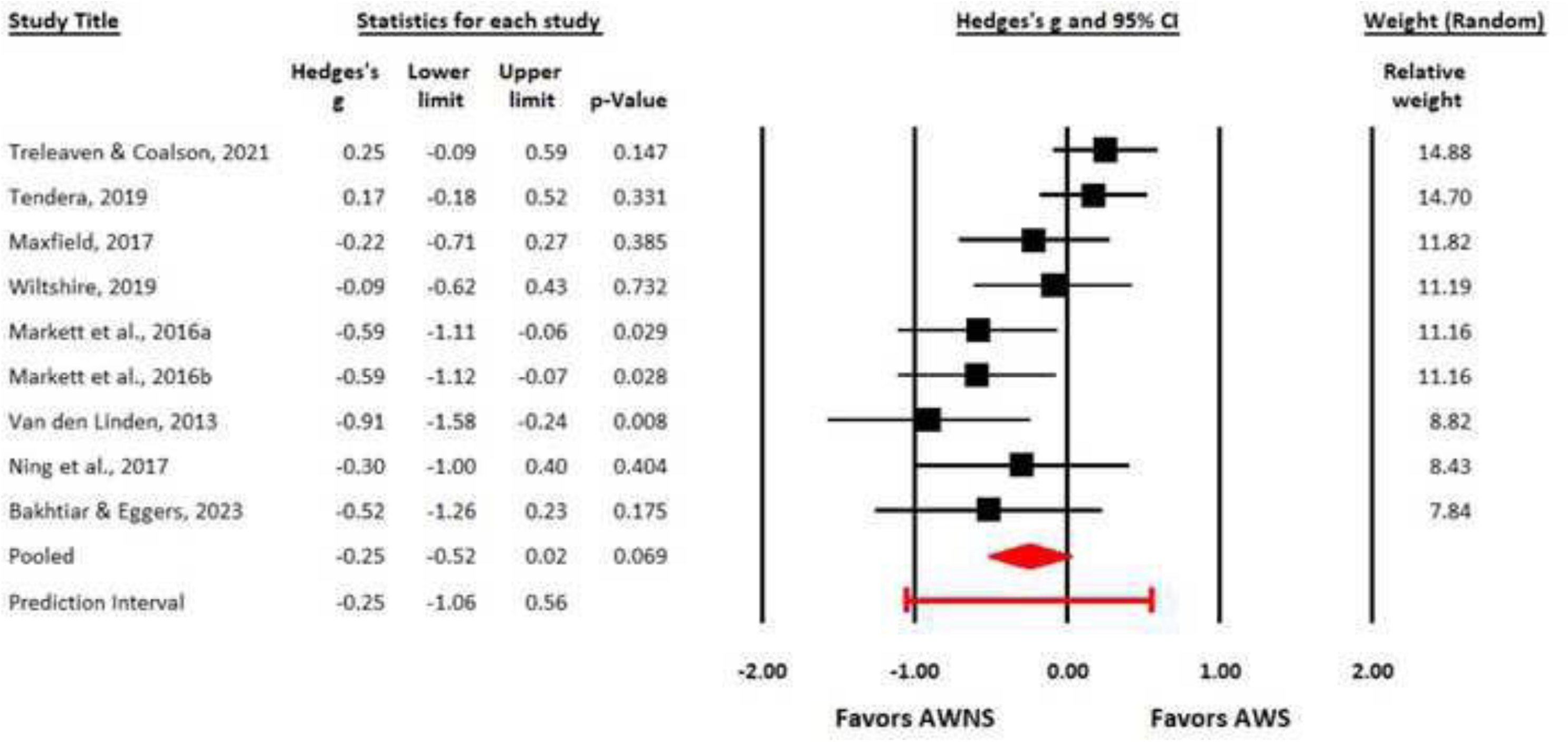
Forest plot for the mean difference in behavioral/motor inhibition control skills between adults who do (AWS) and do not stutter (AWNS). Squares represent effect size per study, with size proportional to its weight in the analysis. Horizontal lines indicate 95% confidence intervals (CIs) per study. The red diamond represents the overall effect size and corresponding 95% CI. The red prediction interval reflects the expected range within which the true effect sizes of future studies fall.

A visual inspection of the funnel plot (see Figure 11) indicated potential evidence of publication bias and the Duval and Tweedie’s trim and fill method estimated that at least four effect sizes with positive Hedges’ *g*-values would be necessary to make the funnel symmetrical (adjusted effect size = 0.03, 95% CI [−0.26, 0.33]). Furthermore, analysis of heterogeneity revealed significant variation in effect sizes across the studies, *Q* = 20.09, df = 8, *p* = .01. Based on the *I^2^* statistic, approximately 60% of the variance in observed effects reflects variance in true effects rather than sampling error. Based on the prediction interval, the true effect size in 95% of all comparable future studies will fall between −1.06 and 0.56.

**Figure 11.**
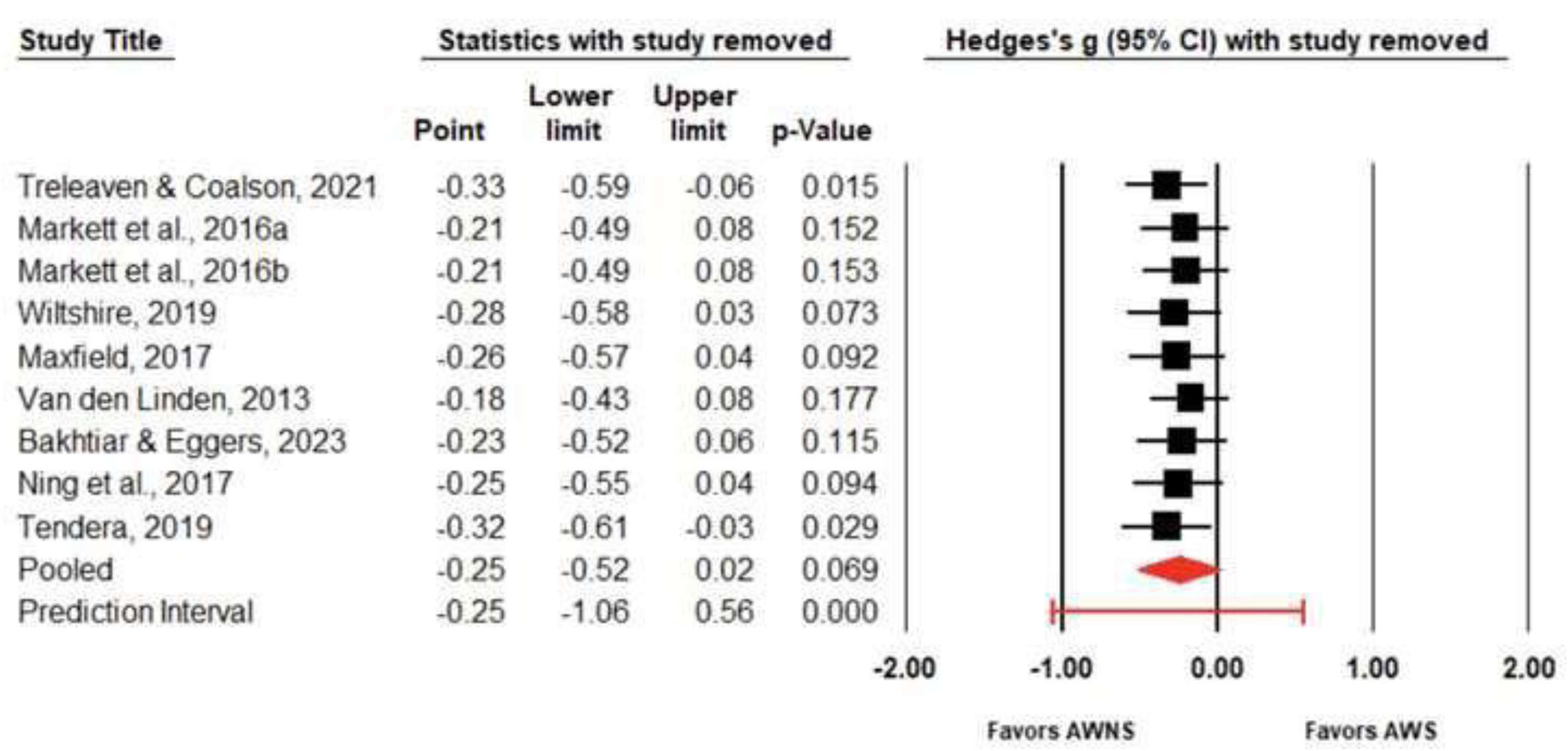
Funnel plot for studies that examined behavioral/motor inhibition skills of adults who do (AWS) and do not stutter(AWNS). The white circles represent individual studies, and white diamonds represent the pooled estimate. Five imputed studies, based on Duval and Tweedie’s trim and fill method, are shown as filled circles, with filled diamonds reflecting the imputed/“adjusted” Hedges’ *g* estimates.

Finally, we performed a sensitivity analysis with the “one-study removed” (see Figure 12) approach. Upon removing the Treleaven and Coalson (2021) or the Tendera (2019) studies, the pooled effect size changed from −0.25 to −0.33 and −0.32, respectively, and became statistically significant, with AWS scoring significantly lower than AWNS in measures of inhibition. Thus, it seems that these two studies had a notable impact on the pooled effect size. Therefore, while the overall trend supports that AWS exhibit poorer inhibitory control than AWNS, the findings highlight the importance of carefully considering the influence of individual studies on the overall effect size.

**Figure 12.**
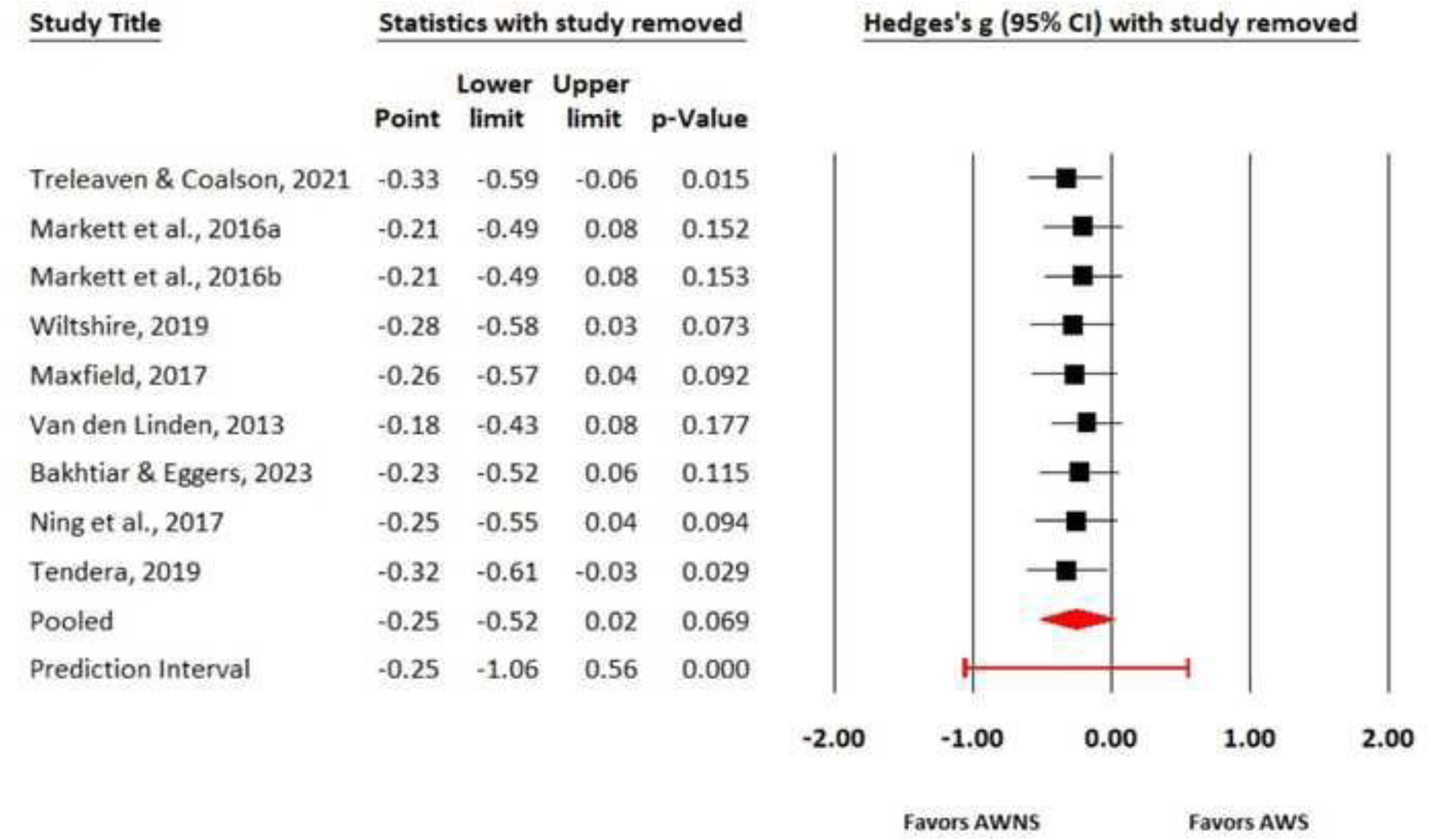
Forest plot for the one-study removed sensitivity analysis. Each line/row displays the pooled effect size and its 95% confidence interval when the respective study was excluded from the analysis.

#### Oculomotor Inhibition

During the timeframe for this review, only one study (Gkalitsiou et al., 2020) examined oculomotor inhibition measures between AWS and AWNS. Given that a minimum of two studies is technically required to conduct a meaningful meta-analysis (Borenstein et al., 2021; Valentine et al., 2010), a descriptive summary of the study is provided as follows: Gkalitsiou et al. (2020) examined oculomotor inhibition in 17 AWS and 17 AWNS using the antisaccade task. On this computer-based task, participants were required to inhibit their prepotent instinct to look at a sudden target stimulus (prosaccade) and instead make a voluntary eye movement in the opposite direction of the stimulus (antisaccade). The authors did not find significant differences in accuracy or reaction time in the antisaccade trials between AWS and AWNS, suggesting a lack of deficit in oculomotor inhibition in AWS.

#### Cognitive Inhibition

No study was identified that examined cognitive inhibition measures between AWS and AWNS.

#### Overall Inhibition

Similar to the overall working memory analysis, we calculated an overall inhibition effect size by combining the four studies on interference control, the eight studies on behavioral/motor inhibition, and the study on oculomotor inhibition (Gkalitsiou et al., 2020). The Markett et al. (2016) study included two independent samples, so it provided two separate effect size estimates to the analysis. The results of the random effects estimate of the overall mean effect size revealed a weighted small and non-significant effect size of Hedges’ *g* = -.10 (SE = .11, 95% CI [-.31, .12]). These results suggest that AWS and AWNS do not differ significantly in inhibition (see Figure 13). A visual inspection of the funnel plot and the Duval and Tweedie’s trim and fill method did not reveal any evidence of publication bias.

**Figure 13.**
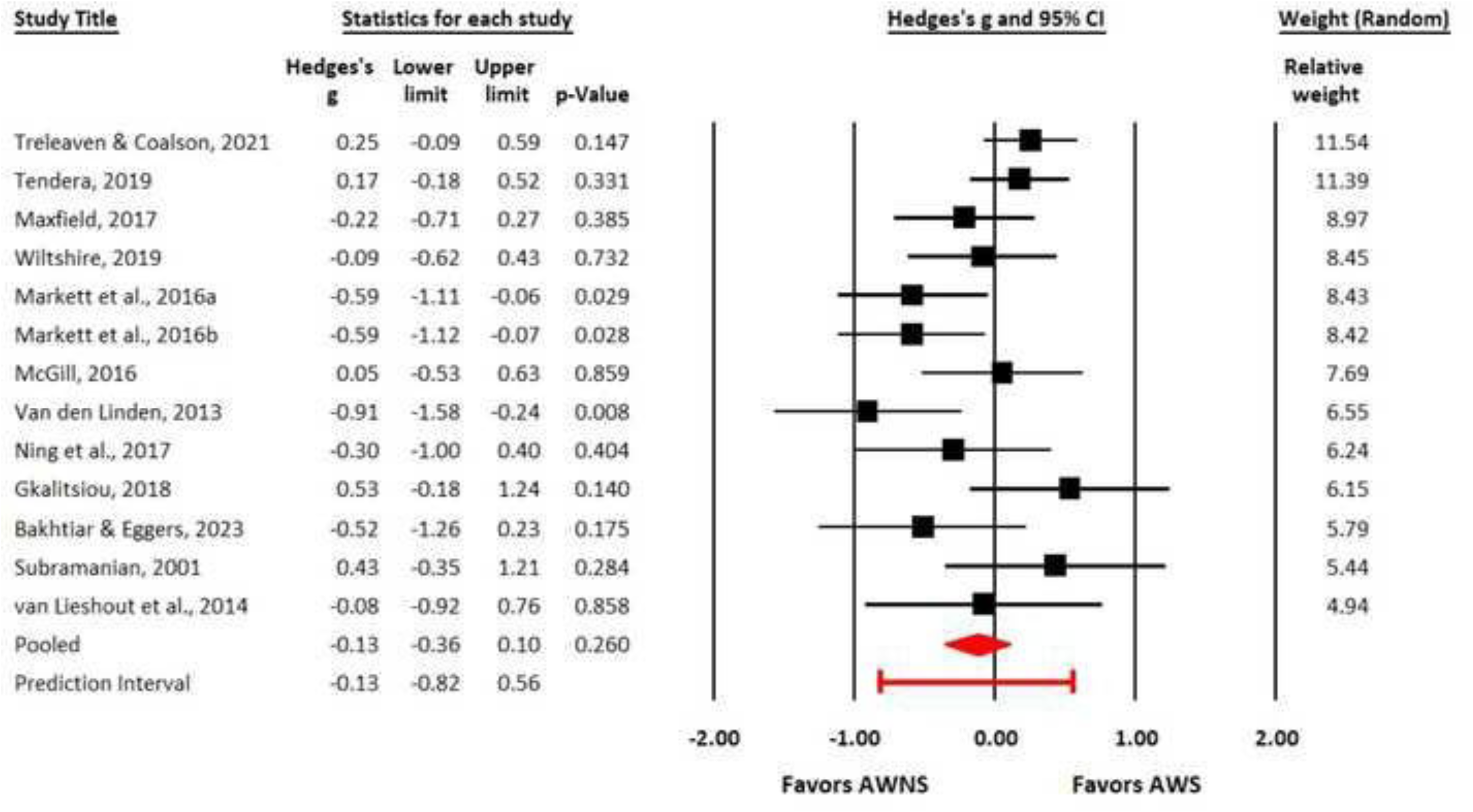
Forest plot for the mean difference in overall inhibition between adults who do (AWS) and do not stutter (AWNS). Squares represent effect size per study, with size proportional to its weight in the analysis. Horizontal lines indicate 95% confidence intervals (CIs) per study. The red diamond represents the overall effect size and corresponding 95% CI. The red prediction interval reflects the expected range within which the true effect sizes of future studies fall.

Analysis of heterogeneity revealed significant variation in effect sizes across the studies, *Q* = 27.03, df = 13, *p* = .01. Based on the *I^2^* statistic, approximately 52% of the variance in observed effects reflects variance in true effects rather than sampling error. This heterogeneity is not surprising given the different tasks/measures included in the analysis. Based on the prediction interval, the true effect size in 95% of all comparable future studies will fall between −0.75 and 0.56. Finally, the “one-study removed” sensitivity analysis indicated that the average effect size remained small and not statistically significant with any one study removed.

### Cognitive Flexibility

To date, three studies (i.e., Doneva et al., 2018; Gkalitsiou, 2018; Markett et al., 2016) that examined cognitive flexibility in AWS and AWNS met the inclusion criteria for the present study.

However, as may be recalled, while a minimum of two studies is required to conduct a meta-analysis (Borenstein et al., 2021), three studies may likewise also be insufficient to assess meaningful heterogeneity and perform a sensitivity analysis. Thus, we provide a descriptive summary of these studies as follows: Markett et al. (2016) used the Category Switch Task (Mayr & Kliegl, 2000) to examine the cognitive flexibility skills of 28 AWS and 28 AWNS, and found no significant differences in the accuracy (*p* = 0.14) or reaction time (*p* = 0.48) between the two groups. Subsequently, Doneva et al. (2018) utilized the Visual Elevator task sub-test from the Test of Everyday Attention (Robertson et al., 1994) to investigate attentional switching/cognitive flexibility in 50 AWS and 50 AWNS. They also found no significant differences in the estimated time per switch between the two groups, *p* = 0.72. Finally, Gkalitsiou (2018) also found no significant differences between the two groups after administering the NIH-Dimensional Change Card Sorting (DCCS) Test to 15 AWS and 15 AWNS, *p* = 0.13. Overall, the current evidence suggests that there are no significant differences between AWS and AWNS in cognitive flexibility skills; however, such findings should be interpreted with caution given the paucity of studies in this core domain of executive function. In addition to the variation in sample sizes, there are important differences in the tasks across the studies. Whereas the Test of Everyday Attention has normative data for adults and is typically used in clinical settings, the Category Switch Task is experimental, with unknown ecological validity. On the other hand, the NIH-Dimensional Change Card Sorting is adaptable to different age groups and has moderate ecological validity (Zelazo et al., 2015). Given the methodological variations, further studies are needed to provide additional insight into the nature of cognitive flexibility skills in AWS.

## Discussion

The purpose of this study was to compare the performance of AWS with that of AWNS on behavioral tasks of executive function in three specific subdomains: working memory, inhibition, and cognitive flexibility. A review of published studies and dissertations was systematically conducted, followed by a meta-analysis of the quantitative measures in each subdomain across the eligible studies. Results indicate that AWS, as a group, performed significantly lower than AWNS on tasks of working memory. However, effect size differences between the two groups were within 1 *SD*, suggesting that, on average, AWS demonstrate relative, rather than clinical, differences in working memory when compared to AWNS. No significant difference in inhibition skills was identified across studies. Cognitive flexibility could not be assessed due to the low number of published studies meeting eligibility criteria for meaningful analysis. These findings support the notion that, like children who stutter, AWS demonstrate subtle yet significant difficulties in at least one subdomain of executive function skills.

### Working Memory

Across tasks, overall working memory abilities were significantly lower for AWS than AWNS (*n* = 31 studies; Hedges’ *g* = -.39, moderate effect size, *p* < .001). In terms of working memory tasks, AWS were found to be significantly less accurate on tasks of nonword repetition (*n* = 15 studies; Hedges’ *g* = - .57, moderate effect size, *p* < .001), forward digit span (*n* = 18 studies; Hedges’ *g* = -.23, small effect size, *p* = .018), and backward digit span (*n* = 11 studies; Hedges’ *g* = -.38, moderate effect size, *p* = .004). Sensitivity analyses confirmed these findings were present after conservative, step-down re-analysis. A final analysis of OSpan revealed potential group differences favoring AWNS (Hedges’ *g* = -.49, small-to-moderate effect size, *p* = .017); however, due to the limited number of studies (*n* = 4), results for OSpan should be interpreted with caution. Although no publication bias was confirmed for any specific task (i.e., nonword repetition, forward or backward digit span, OSpan), such potential was identified for analysis of overall working memory and behavioral inhibition.

The findings that AWS exhibited lower accuracy on nonword repetition and forward digit span tasks are consistent with the meta-analytical review of children who stutter by Ofoe et al. (2018). Like Ofoe et al., the between-group difference in accuracy on the forward span tasks was less pronounced than the differences on the nonword repetition tasks. The magnitude of group differences was also similar for AWS in the present study and children who stutter in the Ofoe et al. study, in that accuracy did not exceed 1 *SD* of the mean for either age group. Based on the similarity between studies, one may apply similar interpretations across age groups in each study. First, given the larger effect size for nonword repetition relative span tasks, differences in working memory in AWS appear to be more evident in the absence of long-term lexical knowledge. Second, given the modest (yet significant) effect size favoring AWNS across tasks, difficulties in working memory in AWS are likely not clinically meaningful and do not reflect a clinically disordered system. Instead, these differences may reflect a working memory system that is less robust and/or more vulnerable to breakdown as demands increase, and particularly when access to long-term lexical knowledge is limited.

However, unlike Ofoe et al. (2018), the inclusion of standardized measures (e.g., nonword repetition: CTOPP; CTOPP-2; Wagner et al., 1999; 2013) to assess nonword repetition (10 of the 15 studies) for adult participants provided an opportunity to examine how differences in phonological working memory in each group compare to the general population. Review of the standard scores of AWS across the studies further supports the notion that difficulties in phonological working memory relative to AWNS, although significant, are subtle. For example, of the 10 studies that included a standardized measure of nonword repetition, only one study reported mean standard scores for AWS below 2 *SD* (i.e., Elsherif et al., 2021, *M* standard score = 7). Moreover, of the four studies that provided individual participant CTOPP scores for nonword repetition, a similar proportion of participants from each group performed within typical limits (+/- 2 *SD*) compared to the general population (i.e., 70% of 50 AWS; 71% of 55 AWNS). That being said, the distribution of group scores differed. Specifically, 22% of the 30% of AWS who did not score within the typical limits, scored 2 *SD* below the population mean, compared to only 5% of AWNS (out of 29% of AWNS who did not score within the typical limits).

Although the CTOPP does not provide scores by syllable length, low overall scores typically indicate that individuals reached ceiling at relatively short nonword lengths, suggesting difficulties even with items as short as 3 syllables. Thus, our finding that a greater percentage of AWS than AWNS scored 2 SD below the population mean suggests that a sizable subset of AWS in these studies was less accurate than the general population during initial encoding and storage of shorter phonological stimuli (e.g., < 3 syllables) that likely do not require rehearsal prior to repetition. Difficulty with syllables as short as 2-to 3-syllables, which would be expected to improve with age, is surprising for the adult speakers and suggests that, in addition to difficulties in phonological rehearsal, difficulties with initial storage and encoding observed for younger children by Ofoe et al. (2018) at 2-syllables does not necessarily disappear with age for a non-trivial number of AWS.

### Inhibition

Across inhibition tasks, AWS demonstrated overall comparable performance compared to AWNS (Hedges’ *g* = -.10, *p* = .37). In terms of interference control, AWS performed similarly to AWNS (Hedges’ *g* = .19, *p* = .23) during tasks that required ignoring distracting stimuli (i.e., Flanker tasks, Stroop tasks, Continuous Processing tasks), although the number of studies identified and eligible for analysis was low (*n* = 4). In terms of behavioral/motor inhibition, AWS performed similarly to AWNS (Hedges’ *g* = -.25, small effect size; *p* = .07) during tasks that require suppression of a dominant movement or response (i.e., SST, Go-NoGo). An insufficient number of studies investigating oculomotor inhibition between AWS and AWNS were identified for analysis, and no studies on cognitive inhibition in AWS and AWNS were identified for analysis.

In terms of cross-study comparison with children who stutter, Ofoe et al. (2018) reported that significant differences in behavioral inhibition between children who do and do not stutter were restricted to parent-report measures. Ofoe et al. reported no group differences on controlled, experimental tasks based on the limited number of studies that incorporated behavioral tasks of inhibition (e.g., Go-NoGo, SST, Grass-Snow/Baa-Meow) similar to those included in this meta-analysis, although the number of child studies that included such tasks was low (*n* = 3). As noted by Ofoe et al., laboratory administration of inhibition tasks may be too challenging for children, particularly complex tasks such as the Grass-Snow/Baa-Meow, wherein participants are required to not just withhold response, but interrupt an in-progress response and/or respond in the presence of conflicting information. Findings from the present study suggest that, even for adult participants who are equipped to complete challenging experimental tasks across a larger number of studies (*n* = 13), participants who stutter performed comparably to non-stuttering participants during behavioral measures of inhibition.

It is important to note that sensitivity analysis indicated that upon removal of two studies (i.e., Tendera, 2019; Treleaven & Coalson, 2021), AWS performed significantly more poorly than AWNS. Both studies compared stop-signal responses between AWS and AWNS, and unlike the other inhibition studies included in our analyses, both asked participants to provide manual as well as vocal responses. It is possible that averaging across two distinct response modalities resulted in atypically faster overall responses for AWS. However, notable between-group disparities were reported for only one study and in one modality – Treleaven and Coalson (2021, manual responses, difference in *M*s = 66 ms) – with all other between-group differences being similar across studies per modality (differences in *M*s ranged from 4 to 9 ms). No other systematic factor shared by these studies (e.g., presentation modality, stimuli complexity, gender balance, stop-signal calculation procedures) was identified to potentially account for the stronger inhibition by the AWS observed in Tendera (2019) and Treleaven and Coalson (2021).

Although these studies fell outside the funnel plot in Figure 11, the standardized residuals for these studies (1.28, 1.52, respectively) did not exceed the standard threshold for removal (>3) in absolute magnitude (Hedges & Olkin, 1985; Viechtbauer & Cheung, 2010). Furthermore, the potential publication bias (*Q* = 20.09; *p* = .01) limits confidence in labelling the group performance of these specific AWS cohorts as outliers. Given the wide range in prediction intervals (−1.06 to .56), it is difficult to determine the true effect size due to heterogeneity among the groups. As such, the present outcomes suggest that while AWS may exhibit inhibition differences relative to AWNS upon inclusion of additional future studies, current data only provide support to the observation that inhibition skills are heterogeneous in AWS and, across studies, statistically comparable AWNS.

A further consideration when interpreting findings related to inhibition measures is the expected methodological differences across studies, such as variation in stimulus presentation (i.e., auditory, visual), stimulus type (i.e., semantic, non-semantic), response modalities (i.e., manual, vocal), and outcome measures. For example, Maxfield (2017) included a combination of semantic and phonemic cues during stimulus presentation. In these cases, we opted to average across conditions rather than further subdivide analyses. In other cases, decisions regarding outcome similarity by the authors were required to bolster comparison across studies. For example, Tendera (2019) used a standard stop-signal task, but chose a less common outcome measure (i.e., point of subjective equality, or PSE) rather than SSRT. We elected to include these findings related to ‘reactive’ interference, rather than ‘proactive interference’, as the underlying processes of reactive interference were akin (but not identical to) standard SSRT outcomes used in other eligible studies. We acknowledge that these decisions, which were made to maintain statistical power and facilitate cross-study comparison, may obscure a more nuanced interpretation of data and potential inhibition differences in AWS. In sum, the present findings did not indicate statistical differences in inhibition in AWS, but the low number of studies, potential publication bias for behavioral inhibition, and heterogeneity across studies provide a window for discovery in future studies examining AWS inhibition skills.

### Cognitive Flexibility

As may be recalled, only three studies have reported information on cognitive flexibility in AWS and AWNS within the timeframe and parameters of the systematic review. However, it is noteworthy that none of these three studies reported significant differences in cognitive flexibility between AWS and AWNS. Ofoe et al. (2018) also reported a similarly low number of studies investigating cognitive flexibility in children who stutter (*n* = 2, search ended December 2016), which precluded meaningful analysis. It should be noted that a number of studies examining differences in cognitive flexibility between CWS and CWNS (e.g., Anderson et al., 2020; Eichorn & Pirutinsky, 2021; Ntourou et al., 2018; Paphiti et al., 2022; Paphiti & Eggers, 2022) have been conducted since Ofoe et al.’s systematic review. Collectively, and similar to working memory and inhibition, findings from these more recent studies indicate subtle challenges in cognitive flexibility for younger individuals who stutter. Given the inter-relationship between each component of executive function skills (see Davidson et al., 2006) and the growth of data in cognitive flexibility in younger children who stutter, further investigation of this component of executive function in AWS and AWNS is warranted.

### Executive Function and Stuttering Persistence

The current findings could be interpreted through the lens of the *Executive Function Model of Developmental Stuttering* (Anderson & Ofoe, 2019), which posits that stuttering could directly or indirectly tax executive function skills or vice versa. Thus, it is possible that a lifetime of frequent fluency breakdowns could partially, and over time, contribute to weaknesses in executive function skills in AWS. According to Anderson and Ofoe (2019), weaknesses in the components of executive function and aspects of attention could disrupt the development and maintenance of robust linguistic representations in the mental lexicon (e.g., Gathercole, 2006). Subsequently, attempts to retrieve the less stable representations from the mental lexicon for production may interfere with the flow of speech. Thus, considering the findings from the present study, as well as those of Ofoe et al. (2018) and Doneva (2020) on the differences in selective/focus attention between AWS and AWNS, it is possible that aspects of attention and executive function may contribute to the development and/or persistence of stuttering.

Furthermore, regarding the components of executive function, given the similarity in performance on working memory tasks between CWS and AWS relative to fluent peers, it is tempting to conclude that executive function, particularly phonological working memory, is related to stuttering persistence. Findings extend differences in phonological memory between young children who stutter (Ofoe et al., 2018) and between young children who persist and recover from stuttering (Spencer & Weber-Fox, 2014; Walsh et al., 2021). Collectively, these findings suggest that weaknesses in phonological working memory skills present early in life in children who stutter are also present in adults whose stuttering persisted, and therefore provide indirect support of the link between phonological working memory and stuttering persistence (e.g., Walsh et al., 2018). These cross-sectional data, however, cannot directly speak to the trajectory of these individual participants. For example, based on these data, we cannot conclude that participants who performed poorly as adults in these studies also struggled at younger ages. We also cannot account for the executive function skills of those children who no longer stutter but who may, hypothetically, have exhibited similarly low executive function skills as a child. Further comparison of AWS and adults who recovered from stuttering during childhood would be required to confirm the potential link between persistence and phonological working memory. Additional considerations—such as concomitant phonological or language issues, as well as treatment history — would also warrant attention as potential mediating factors. Together, a more comprehensive account would be to include all studies, across ages, and account for persistence rather than, or in addition to, age.

### Executive Function Skills, Psychosocial Functioning, and Quality of Life in AWS

As previously noted, executive function skills are linked to psychosocial development, mental health, and overall quality of life across the lifespan (Brown & Landgraf, 2010; Miller et al., 2011).

Theoretically, weaknesses in aspects of attention and executive function skills could compromise the efficient planning and execution of speech-language production in people who stutter. In the event of an increased cognitive load, these weaknesses may lead to disruptions in the fluency of their speech-language production. Conversely, people who stutter may attempt to counteract the frequent speech breakdowns by excessively relying on their diminished executive function and attentional resources to produce fluent speech, which may further weaken their cognitive skills and impact other processes, including socioemotional skills (see Anderson & Ofoe, 2019; Choo et al., 2020). Given that speech and communication skills are essential for social and emotional functioning (van Barreveld et al., 2025; Rautakoski et al., 2021), disruptions in the speech fluency of individuals who stutter may increase their risk of developing speech-related anxiety (Blumgart et al., 2010; Craig & Tran, 2006; Gabel et al., 2002; Iverach et al., 2011), which can impact the overall effectiveness of verbal communication in social settings. Over time, the vulnerabilities and disruptions in the flow of speech during social interactions may cause or exacerbate feelings of shame, embarrassment, and lower self-esteem, which may lead to withdrawal or avoidance of social situations and impact their education, career choices, and overall quality of life (Craig et al., 2009; Daniels & Gabel, 2004; Hayhow et al., 2002; Langevin & Prasad, 2012; Koedoot et al., 2011). Beyond the potential relationship between speech fluency and executive function, it is possible that negative experiences with stuttering may be mediated by executive function. For example, AWS have reported increased repetitive negative thinking related to previous communication experiences (Tichenor & Yaruss, 2020) - a process linked to one’s (in)efficiency when updating working memory in AWNS (Zetsche et al., 2018). However, Coalson et al. (2025) found that skills related to updating the content of working memory were not associated with increased rumination in AWS.

Although further investigation of the link between executive function and quality of life in AWS has yet to reach consensus, there is clear potential to explore this hypothesis in future studies.

### Limitations and Future Studies

There are a number of limitations that warrant consideration. First, the age of inclusion was typically 18 years, which may be too young to capture significant milestones in executive function (e.g., Korzeniowski et al., 2021). Second, as is common in meta-analyses, these findings represent group trends rather than individual differences in executive function skills. That is, these outcomes should not be seen as prescriptive to all individuals who stutter, but rather as overall differences that inform factors that contribute to, and inform, stuttering as a condition. We also acknowledge that the measures used in these studies are, for the most part, broad metrics that require a behavioral response from the participants. To maintain a sufficient sample size for robust meta-analyses, studies were grouped by methodology in accordance with the overarching theoretical framework within each domain of executive function (e.g., Baddeley and Hitch’s framework of working memory, Nigg’s taxonomy of inhibition). The outcomes of this study, however, cannot account for group differences attributable to study-specific variations or modifications of tasks (e.g., stimulus presentation, response modality, calculation of outcome measures). Even during the administration of more standardized measures (e.g., the nonword repetition task on CTOPP), outcomes may be influenced by variations in speech rate, strategy use, examiner familiarity, fluent versus disfluent trials, and other factors. Thus, although valuable, we acknowledge that such factors are typically allowed to vary freely across studies and may obscure differences (or lack thereof) obtained through these subtle but important variations.

Furthermore, such behavioral measures, while they are direct measures of the participants’ performance, may not be sensitive enough to capture underlying differences more apt for examination by neurophysiological measures. Additionally, future studies may consider incorporating and reporting self-report measures, which involve reflections about thoughts, feelings, and behavior in different situations, in addition to the behavioral measures to provide a more comprehensive perspective of executive function skills between AWS and AWNS (see Dang et al., 2020). Furthermore, few studies screened for ADHD in the participants (e.g., Treleaven & Coalson, 2021) despite the intersection between ADHD and stuttering (Tichenor et al., 2021), or addressed co-occurring mental health concerns that may be related to atypical executive function skills (e.g., depression: Zetsche et al., 2018; anxiety: Moran, 2016); future studies may consider incorporating these measures into their study designs.

Finally, it is noteworthy that some publication bias may be present and, relatedly, many of the studies eligible for inclusion in the meta-analysis were unpublished dissertations and theses. To enhance the quality of future meta-analytic investigations and cross-study comparisons, future researchers should strive not only to include the minimum necessary data (e.g., control groups, descriptive demographic data, and required descriptive and statistical information) but also to seek studies that yield data that counter the documented publication and/or confirmation bias.

### Conclusions

The purpose of this study was to conduct a meta-analytic review of the primary components of the executive function skills of AWS relative to AWNS. The present findings suggest that AWS, as a group, differ subtly yet significantly from AWNS in working memory. Differences in inhibition did not reach significance, although outliers and potential publication bias were detected. At present, insufficient data are available to conduct meaningful cross-study comparisons of cognitive flexibility, and future investigations may consider focusing on this executive function subdomain in adults who stutter.

## Acknowledgments

This research was partly supported by a Research Seed Grant from the University of West Georgia’s College of Education awarded to the first author, a grant from the Oklahoma Center for the Advancement of Science and Technology [HR 21-052-2] awarded to the second author, and a grant from the National Institutes of Health (R21DC018109-01A1) awarded to the fourth author.

## Data Availability Statement

All data generated or analyzed during this study are included in this published article (and/or its supplemental material files, if applicable).

## Appendix A. Talker Classification Criteria across Studies that examined Executive Function between Adults Who Do (AWS) and Do Not Stutter (AWNS)

| Study | Group | SSI-3 /-4 | % SLD | Likert SEV Scales | OASES | Self-ID | SLP Judgment | Other | Total criteria |
| --- | --- | --- | --- | --- | --- | --- | --- | --- | --- |
| Baird (1996) | AWS |  | √<sup>s</sup> |  |  |  |  |  | 1 |
|  | AWNS |  |  |  |  |  | √ |  | 1 |
| Bakhtiar & Eggers (2023) | AWS |  | √<sup>c+r</sup> |  |  | √ |  |  | 2 |
|  | AWNS |  |  |  |  | √ |  |  | 1 |
| Bauerly (2012) | AWS |  |  |  |  |  |  |  | NR |
|  | AWNS |  |  |  |  |  |  |  | NR |
| Brocklehurst & Corley (2011)<sup>†</sup> | AWS | √<sup>c+r</sup> | √<sup>r</sup> |  | √ |  | √ | Self-ratings of hierarchical 'fluency difficulty' | 4 |
|  | AWNS |  | √<sup>r</sup> |  | √ |  |  |  | 2 |
| Brown (2015) | AWS |  |  | √ |  |  | √ | Self-reported history of SLDs | 2 |
|  | AWNS |  |  |  |  |  |  |  | NR |
| Byrd et al. (2015) | AWS | √<sup>c+r</sup> |  | √ |  |  | √ |  | 2 |
|  | AWNS |  |  |  |  |  |  |  | NR |
| Byrd et al. (2017) | AWS | √<sup>s</sup> |  | √<sup>s</sup> |  | √ | √ |  | 4 |
|  | AWNS |  |  |  |  |  |  |  | NR |
| Coalson & Byrd (2015a) | AWS |  |  | √<sup>c+r</sup> |  | √ | √ |  | 3 |
|  | AWNS |  |  | √ |  | √ |  |  | 2 |
| Coalson & Byrd (2015b) | AWS |  |  | √<sup>c+r</sup> |  | √ | √ |  | 3 |
|  | AWNS |  |  | √ |  | √ |  |  | 2 |
| Coalson et al. (2017) | AWS | √<sup>c</sup> |  |  |  | √ | √ |  | 3 |
|  | AWNS |  |  |  |  |  |  |  | NR |
| Eichorn et al. (2016)<sup>††</sup> | AWS | √<sup>s</sup> |  |  | √ | √ | √ |  | 4 |
|  | AWNS | √ |  |  |  |  |  |  | 1 |
| Elsherif (2021) | AWS | √<sup>c+tr</sup> |  |  |  | √ |  |  | 2 |
|  | AWNS |  |  |  |  |  |  |  | NR |
| Garnett et al. (2023) | AWS | √<sup>s</sup> |  |  |  |  |  |  | 1 |
|  | AWNS |  |  |  |  |  | √ |  | 1 |
| Gkalitsiou (2018) | AWS | √<sup>s</sup> |  |  |  | √ | √ |  | 3 |
|  | AWNS |  |  |  |  | √ | √ |  | 2 |
| Gkalitsiou et al. (2020) | AWS | √<sup>r</sup> |  |  |  | √ |  |  | 2 |
|  | AWNS |  |  |  |  |  |  |  | NR |
| Haj-Tas (2007) | AWS |  | √<sup>c</sup> |  |  |  |  | Self-rated severity across situations | 1 |
|  | AWNS |  |  |  |  | √ |  |  | 1 |
| Höbler (2023) | AWS | √<sup>c+tr</sup> |  |  |  | √ |  |  | 2 |
|  | AWNS |  |  |  |  |  |  |  | NR |
| Jackson (2012) | AWS |  |  |  |  | √ |  | Support group membership | 1 |
|  | AWNS |  |  |  |  |  |  |  | NR |
| Lescht et al. (2017) | AWS | √<sup>c+tr</sup> |  |  |  |  |  |  | 1 |
|  | AWNS |  |  |  |  |  |  |  | NR |
| Markett et al. (2016) | AWS |  |  |  |  | √ | √ |  | 2 |
|  | AWNS |  |  |  |  | √ |  | No family history | 2 |
| Maxfield (2017) | AWS |  |  | √<sup>c</sup> | √ |  |  |  | 2 |
|  | AWNS |  |  |  |  | √ |  |  | 1 |
| McGill (2016)<sup>†</sup> | AWS |  |  |  |  | √ | √ |  | 2 |
|  | AWNS |  |  |  |  |  |  |  | NR |
| McGill et al. (2016) | AWS | √<sup>s</sup> | √<sup>s</sup> |  |  | √ | √ |  | 4 |
|  | AWNS | √ | √ |  |  | √ |  |  | 3 |
| Mersov et al.<br>(2016) | AWS | √<sup>s</sup> |  |  |  | √ |  |  | 2 |
|  | AWNS |  |  |  |  | √ |  |  | 1 |
| Ning et al.<br>(2017) | AWS | √<sup>s</sup> |  |  |  | √ |  |  | 2 |
|  | AWNS |  |  |  |  | √ |  |  | 1 |
| Sasisekaran<br>(2013) | AWS | √<sup>s</sup> |  |  |  |  | √ |  | 2 |
|  | AWNS |  |  |  |  | √ | √ |  | 2 |
| Sasisekaran &<br>Weisberg<br>(2014) | AWS | √<sup>s</sup> |  |  |  |  |  |  | 1 |
|  | AWNS |  |  |  |  | √ | √ |  | 2 |
| Schwartz &<br>Kotz (2020) | AWS | √<sup>c</sup> |  |  |  |  |  |  | 1 |
|  | AWNS |  |  |  | √ |  |  |  | 1 |
| Smits-<br>Bandstra &<br>Gracco (2015)<br>++ | AWS | √<sup>c+tr</sup> |  |  |  |  | √ |  | 2 |
|  | AWNS |  |  |  |  |  | √ |  | 1 |
| Subramanian<br>(2001) | AWS |  | √<sup>c</sup> |  |  | √ | √ |  | 3 |
|  | AWNS |  | √ |  |  | √ | √ |  | 3 |
| Tendera<br>(2019) | AWS | √<sup>c+tr</sup> |  |  | √ |  |  |  | 2 |
|  | AWNS |  |  |  |  | √ |  |  | 1 |
| Tichenor<br>(2020) | AWS |  |  |  |  | √ |  |  | 1 |
|  | AWNS |  |  |  |  | √ |  |  | 1 |
| Treleaven &<br>Coalson<br>(2021) | AWS | √<sup>s</sup> |  |  |  | √ | √ |  | 3 |
|  | AWNS | √ | √ |  |  | √ |  |  | 3 |
| Vallely (2011) | AWS |  |  | √<sup>s</sup> |  |  |  |  | 1 |
|  | AWNS |  |  |  |  | √ |  |  | 1 |
| Van der<br>Linden (2013) | AWS |  |  |  |  |  | √ |  | 1 |
|  | AWNS |  |  |  |  |  | √ |  | 1 |
| Van Lieshout<br>et al. (2014) | AWS | √<sup>c+tr</sup> |  |  |  |  | √ |  | 2 |
|  | AWNS |  |  |  |  | √ | √ |  | 2 |
| Vincent (2012) | AWS | √<sup>c</sup> |  |  |  | √ |  |  | 2 |
|  | AWNS | √ | √ |  | √ |  |  |  | 3 |
| Webster (1990) | AWS |  |  |  | √ | √ |  | Self-reported severity overall | 2 |
|  | AWNS |  |  |  |  |  |  |  | NR |
| Wiltshire (2019) | AWS | √<sup>s</sup> |  |  |  |  |  |  | 1 |
|  | AWNS |  |  |  |  |  |  |  | NR |
| Yang et al. (2018) | AWS | √<sup>s</sup> |  |  |  | √ |  |  | 2 |
|  | AWNS |  |  |  |  |  |  |  | NR |
| Total | AWS | 24 | 5 | 9 | 4 | 24 | 19 | 0 |  |
|  | AWNS | 3 | 5 | 3 | 3 | 19 | 9 | 0 |  |

*Note.* AWS: adults who stutter; AWNS: adults who do not stutter; SSI-3/-4: Stuttering Severity Instrument – 3^rd^ Edition [Riley, 1994] and 4^th^ Edition [Riley, 2009]); %SLD: percent of stuttering-like disfluencies within speech sample(s); OASES: Overall Assessment of the Speaker’s Experience of Stuttering (Yaruss & Quesal, 2010); SLP: speech-language pathologist; ID: identification; SLP judgment includes in-experiment confirmation and prior diagnosis and/or treatment by a licensed SLP. NR: criteria not reported

^c^Conversational sample

^r^Reading sample

^s^Speech sample (unspecified)

^†^Groups differentiated by significant difference on OASES-Section IIIA

^††^Groups differentiated by significant differences on SSI-4 raw score

## Appendix B. The date and search strategy for the study

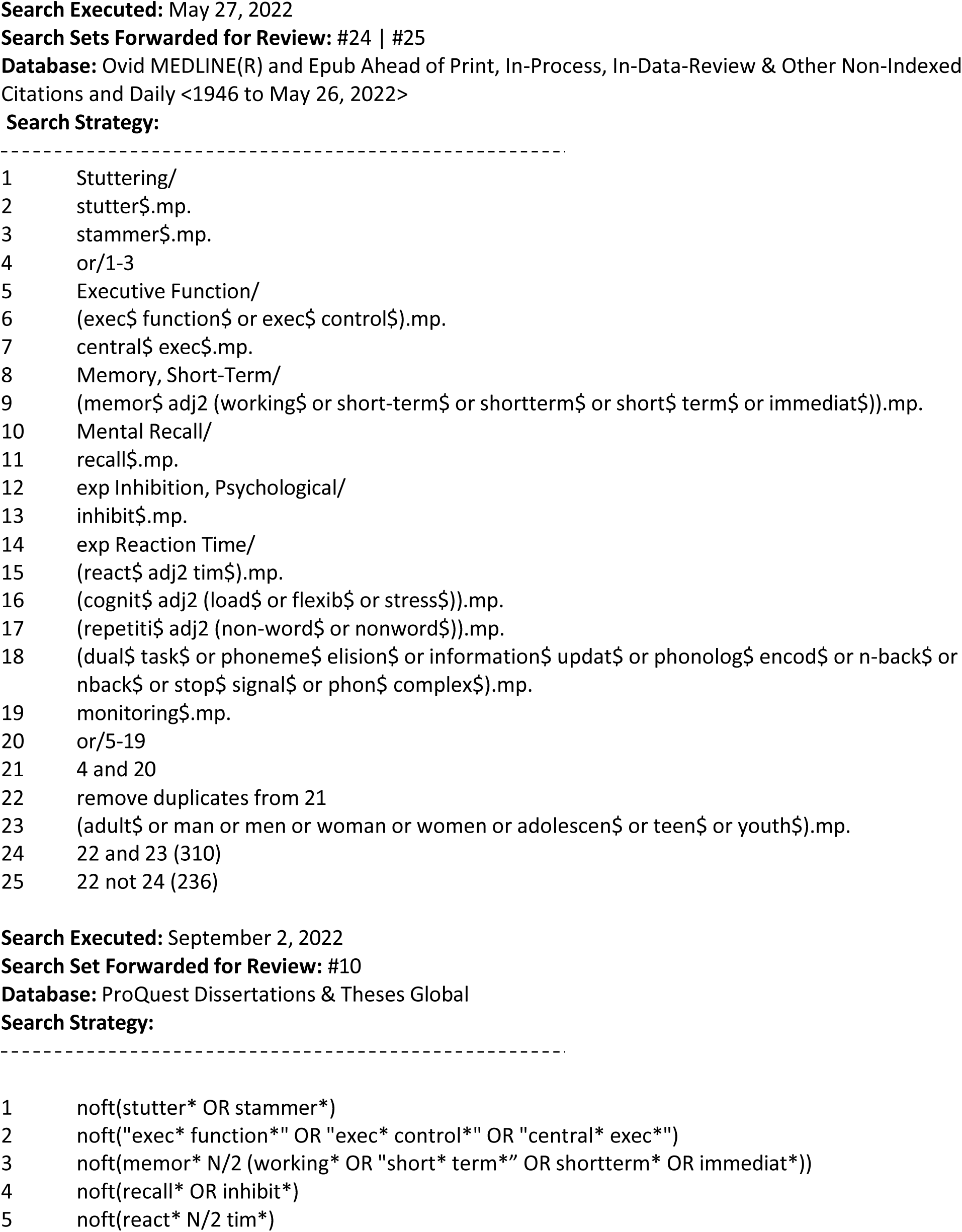
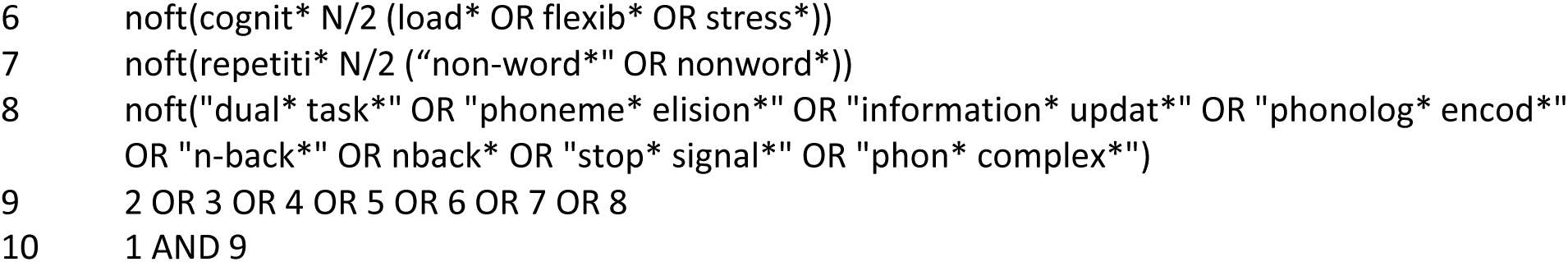

## Appendix C. A Glossary of the Meta-analysis Terms Used to Examine Executive Function between Adults Who Do (AWS) and Do Not Stutter (AWNS)

**Appendix C.** A Glossary of The Meta-analysis Terms Used in The Current Study.

| Term | Meaning |
| --- | --- |
| Hedges' *g* | An effect size measure that expresses the standardized mean difference between two groups, adjusted for small sample size bias. |
| Cochrane Q (Q statistic) | A test of heterogeneity in meta-analysis that assesses whether the variability in effect sizes across studies is greater than would be expected by chance, indicating true differences rather than random sampling error. |
| *I*<sup>2</sup> Index | A statistic that specifies the degree of actual variation rather than random chance among studies in a meta-analysis. |
| Publication bias | The systematic error that occurs when published literature overrepresents studies with significant or favorable results, leading to a biased estimation of the true effect. |
| Funnel plot | Visual tool assessing the possible existence of publication bias. |
| Duval and Tweedie's trim and fill method | An analytic technique that tests for funnel plot asymmetry and imputes missing studies to evaluate the impact of publication bias. |
| Sensitivity analysis | A rigorous method used to examine whether the results or direction of effects are unduly influenced by specific studies. |
| Egger's regression | A linear regression test used to examine the potential publication bias via a funnel plot asymmetry. |
| Random effects model | A type of meta-analysis model that considers the actual effect size variation from study to study due to population differences, rather than assuming that each study shares the same effect size due to samples drawn from the same population. |
| "One study removed" analysis. | A meta-analytic technique where the overall effect size is recalculated repeatedly, each time omitting a single study, to assess whether results are disproportionately influenced by any one study. |

## Footnotes

1 Some researchers associate interference control with attention (Friedman & Miyake, 2004). However, attention as a cognitive construct was not considered in the present study [see Doneva (2020) for a meta-analytic review of attention skills in AWS and AWNS].

2 Behavioral/motor inhibition tasks can be further categorized into simple and complex, with the Go-NoGo and stop-signal tasks classified as simple response inhibition tasks as these tasks do not require a subdominant response (Garon et al., 2008).

